# Utility of genome sequencing and group-enrichment to support splice variant interpretation in Marfan syndrome

**DOI:** 10.1101/2025.02.18.25321172

**Authors:** Susan Walker, David J. Bunyan, Huw B. Thomas, Yesim Kesim, Christopher J. Kershaw, John Holloway, Htoo Wai, Michael Day, Cassandra L. Smith, Gareth Hawkes, Andrew R. Wood, Michael N. Weedon, Ed Blair, Stephanie L. Curtis, Catherine Fielden, Julie Evans, Rebecca Whittington, Sarah F. Smithson, Helen Cox, Paul Clift, Meriel McEntagart, Matina Prapa, Suzanne Alsters, Deborah Morris-Rosendahl, John Dean, Patrick J. Morrison, Abhijit Dixit, Ajoy Sarkar, Katrina Prescott, Leila Amel Riazat Kesh, Riya Tharakan, Claire Turner, Sian Ellard, Charles Shaw-Smith, James Fasham, Virginia Clowes, Simon Holden, Suresh Somarathi, Catherine Mercer, Ian Berry, Raymond T. O’Keefe, Siddharth Banka, Diana Baralle, N. Simon Thomas, Emma L. Baple, Jenny C. Taylor, Alistair T. Pagnamenta

## Abstract

**Purpose:** To quantify the impact of non-canonical *FBN1* splice site variants in undiagnosed Marfan syndrome (MFS), a connective tissue disorder associated with skeletal abnormalities and Familial Thoracic Aortic Aneurysm Disease (FTAAD).

**Methods:** A systematic analysis of ultra-rare *FBN1* variants was performed using genome sequencing data from the 100,000 Genomes Project. Variants were annotated with SpliceAI and the significance of enrichment among individuals with FTAAD was assessed using Fisher’s exact test. Experimental validation utilised RNAseq, RT-PCR, minigene constructs and replication analysis was with data from UK Biobank.

**Results:** Using aggregate data for 78,195 individuals, we identified 13,864 singleton SNVs in *FBN1* of which 21 were predicted to impact splicing (SpliceAI >0.5). Incidence of candidate splice variants in individuals recruited with FTAAD (9/703) was significantly elevated compared with that seen in non-FTAAD participants (12/77,492; OR=84, *p*=9.7×10^−14^). Additional analysis uncovered a further 14 families harbouring 11 different *FBN1* splice variants. A total of 20 candidate splice variants in 23 families were identified, of which 70% lay beyond the ±8 splice regions. RNA testing confirmed the predicted splice aberration in 14/20 and for 9/20, pseudoexonization was the likely splicing-anomaly.

**Conclusion:** Our findings indicate that non-canonical splice variants may account for ~3% of families with undiagnosed FTAAD, highlighting the importance of incorporating analysis of introns and confirmatory RNA testing into genetic testing for MFS.

## INTRODUCTION

Marfan syndrome (MFS; MIM #154700) is a connective tissue disorder first described in 1896, with variable penetrance/expressivity, an estimated incidence of 2-3 in 10,000 individuals and an average age at diagnosis in the early 20s.^1–3^ In addition to a number of typical skeletal features, aneurysm of the thoracic aorta is a cardinal feature, which can in the absence of surgery progress to sudden aortic dissection and death. The “Ghent nosology” which primarily comprises a set of cardiovascular, ocular and skeletal manifestations was developed to facilitate clinical diagnosis of this syndrome and improve patient management/counselling.^4^ Although the first pathogenic variants in *FBN1* (HGNC:3603) described in MFS patients >30 years ago were *de novo* in origin^5^, due to variable effects on fecundity, inherited pathogenic variants have also been found cosegregating across large multigenerational pedigrees.^6,7^ *FBN1* is a large gene spread over 237kb, with 66 exons of which 65 are coding. The gene encodes fibrillin 1, a glycoprotein that is localised to the extracellular matrix. Pathogenic variants reported to date frequently include those resulting in premature termination codons (PTCs; including those in the last exon), in-frame indels and missense variants, particularly those disrupting cysteine residues that impact on the formation of structurally important disulphide bonds.^8–10^ Geleophysic and acromicric dysplasia (MIM #614185 and #102370), conditions characterized by severe short stature, short extremities and stiff joints, have also been linked to variants in *FBN1* but with pathogenic variants residing in exons 41 and 42.^11^

Using conventional methods, rare pathogenic or likely-pathogenic *FBN1* variants can be identified in >90% of individuals with clinically diagnosed MFS.^12,13^ Families without a molecular diagnosis following clinical testing may be explained by poor phenotypic characterisation, the presence of variants in other gene(s) linked to related conditions^13^, or cryptic variants in *FBN1*. Recently, the identification of structural variants, including inversions^14,15^, balanced translocations^16^ and complex interchromosomal insertions^17^, has highlighted the benefits of using genome sequencing data and analysing intronic sequence of *FBN1* for genetically undiagnosed cases of MFS.

Genome sequencing also enables detection of cryptic splice variants that lie beyond ±8bp into intronic sequence, which often remain undetected by current clinical testing procedures. Although many pathogenic variants in the literature result in aberrant splicing, reports of non-canonical splice variants in *FBN1* have mainly been limited to case studies (Table S1) and many of these were detected using RNA-based testing approaches.

In this study, we performed a systematic assessment of genome sequence data from the 100,000 Genomes Project (100kGP) which included >700 individuals recruited with a diagnosis of Familial Thoracic Aortic Aneurysm Disease (FTAAD). Although there are over 30 genes linked to this condition, the diagnostic yield for this set of families is below 10%. We aimed to identify new molecular diagnoses for families with suspected MFS and in parallel assess the utility of genome sequencing in combination with *in silico* splice prediction, for the detection of cryptic variants in *FBN1*.

## MATERIALS AND METHODS

### Bioinformatic extraction/annotation of singleton variants

A systematic analysis of ultrarare *FBN1* variants was performed using an aggregated dataset produced from v10 release of the 100kGP, a nationwide study that aimed to uncover the genetic basis of disease for individuals in whom a diagnosis had not been obtained by standard-of-care testing.^18^ The aggregation included germline data for 78,195 participants from both the rare disease and cancer programme of the 100kGP, with FTAAD, being the 11^th^ most common disease indication (Table S2). This dataset (the “AggV2” file) includes >722 million SNVs and small indels (≤50 bp) and is split into 1,371 genome chunks of approximately equal size. DNA samples were quality controlled and genome sequencing was conducted using a HiSeqX instrument (Illumina) generating 150bp paired-end reads. Alignment was to the GRCh38 assembly. Further information about data processing and the construction of this aggregate data is available in the Supplementary Methods and at https://re-docs.genomicsengland.co.uk/aggv2.

Singleton variants were extracted from the AggV2 file using BCFtools for the entire *FBN1* gene region (chr15:48,408,313-48,645,709, GRCh38). Variant annotation was performed using VEP and HGVS annotation of variants was based on the MANE select transcript (NM_000138.5). Odds Ratios (ORs) and 2-tailed significance levels based on Fisher’s exact test were calculated using RStudio. Additional scrutiny of *in silico* splice predictions and the surrounding landscape of absolute SpliceAI scores was performed using SpliceAI-visual^19^, with the resulting bedGraph files downloaded and imported as a custom track into the UCSC genome browser. The SpliceAI lookup tool (https://spliceailookup.broadinstitute.org) was also used with the max region of effect increased to 500bp. The option to output absolute scores for REF/ALT alleles was utilized. SpliceAI scores are linked to one of four possible consequences: acceptor gain (AG), acceptor loss (AL), donor gain (DG) and donor loss (DL). Unless otherwise stated, just the highest of the four delta scores (range 0-1) is reported, alongside the delta position.^20^

### Secondary analysis involving FTAAD-only data

The October 2024 data release (v19) of the 100kGP includes 72,884 rare disease programme participants. Included in this dataset were 672 families recruited with FTAAD. The FTAAD category was created in the 100kGP to encompass patients with syndromic and non-syndromic forms of aortic dilatation (Supplementary Methods). A number of these families were not available on the GRCh38 genome build in data release v10, i.e. at the time the AggV2 file was created. Thus, these individuals would not have been assessed in our primary analysis. A secondary analysis was therefore performed which included all families recruited to the 100kGP with FTAAD, irrespective of which build had been used for read-mapping. In this analysis, we did not require rare variants to be unique in the 100kGP and used a more sensitive SpliceAI threshold of 0.2. Data was analysed using a range of custom scripts. We also included a family sequenced as part of NHS England’s Genomic Medicine Service (GMS) using similar procedures to those described above, but where the variant had been picked up by the local Genomic Laboratory Hub using a commercial analysis platform (Congenica) to identify intronic substitutions with a SpliceAI score of >0.1.

### RNAseq

RNAseq data are available for 5,546 probands from the 100kGP, of whom 180 were recruited with FTAAD as the primary diagnosis. Further technical details and information about sample prioritisation are available in the Supplementary Methods and elsewhere.^21^ In brief, total RNA was extracted from peripheral whole blood using the PAXgene Blood RNA Kit (Qiagen, UK). Libraries were constructed following the Illumina Stranded Total RNA Prep kit and ligation was with the Ribo-Zero Plus protocol. Sequencing was done on the NovaSeq 6000 using 2×100bp paired-end reads.

### Targeted RNA analysis

For targeted RNA testing, blood samples were collected into PAXgene RNA collection tubes. cDNA preparation, RT-PCR and Sanger sequencing was then carried out as described in the Supplementary Methods and using the primers listed in Table S3. The one exception to this was Family 20, where fibroblasts were cultured in the presence/absence of cycloheximide in order to monitor levels of nonsense-mediated decay (NMD).

### Minigene constructs

Minigene vectors for both wild-type and variant sequences of *FBN1* were assembled by Gibson cloning using the SK3 minigene vector (a derivative of the pSpliceExpress minigene splice reporter vector, gifted by S. Stamm; Addgene, 32485, RRID:Addgene_32485), as previously described.^22^ Human embryonic kidney 293 cells were cultured to 60-75% confluency in 2ml of high-glucose Dulbecco’s modified Eagle’s medium (Gibco), supplemented with 10% foetal bovine serum (Gibco) in a six-well culture plate at 37°C with 5% CO_2_. Cells were transiently transfected with 2μg of either WT or variant minigene constructs using Lipofectamine 2000 (Thermo Fisher Scientific) and the manufacturer’s standard protocol. Following 18-21h incubation, RNA was extracted using 1ml of TRI Reagent® (Invitrogen) per well and further purified using the RNAeasy cleanup kit (QIAGEN) which included a DNase digestion step. cDNA was synthesized from up to 4μg RNA (using an equal amount of RNA for each sample set) by GoScript™ first strand synthesis using the manufacturer’s recommended protocol (Promega). Resulting cDNA was amplified using Q5 Polymerase (New England Biolabs) with the primers listed in Table S3. Finally, PCR products were run on an agarose gel (1-4%) supplemented with SYBR Safe (Thermo Fisher Scientific) for visualization. PCR products were purified using a PureLink™ gel extraction kit (Invitrogen) and sequenced by Sanger sequencing (Eurofins Genomics) to confirm splicing products.

### Analysis of FBN1 splice variants using UK Biobank

UK Biobank (UKB) is a population-based study involving half a million participants from the UK.^23^ Recruitment was between 2006–2010 and participants were aged 40–69 years at time of recruitment. In addition to the comprehensive demographic and health-related measures, short-read genome sequencing was recently performed, as described.^24^ The R package ukbrapR (https://lcpilling.github.io/ukbrapR) for working in the UKB Research Analysis Platform was used to identify UKB participants with the ICD10 codes I71 (Aortic aneurysm and dissection) and Q87.4 (MFS). Variants were annotated using Ensemble VEP v110, as described (https://github.com/drarwood/vep_ukb_aou_docker) and SNVs were extracted from the chromosome 15 file (version of 10th March 2024) in the MANE select transcript for *FBN1* (ENST00000316623.10 / NM_000138.5). To align with the systematic analysis performed on 100kGP data, we filtered for variants with a maximum SpliceAI score of 0.5 or more using precomputed scores and the 50bp analysis window. Ultra-rare variants were identified by filtering using global population allele frequencies from the gnomAD v4 genomes, such that only variants that are singleton variants or else absent in this dataset were retained. Data access was under the approved UKB project “Understanding the role of rare and common genetic variation in human phenotypes”.

## RESULTS

### Identification of putative splice variants in FBN1

To assess the potential contribution of intronic *FBN1* variants in the aetiology of FTADD for families recruited to the 100kGP, we performed a systematic analysis of rare variants with SpliceAI scores indicative of a possible splicing alteration. Initially, the analysis included 78,195 participants of the 100kGP, 703 of whom were recruited with FTAAD, i.e. 0.90% of the total cohort (Table S2). We considered singleton variants (allele count of 1 in the AggV2 dataset) in *FBN1* across all phenotype categories and identified 13,864 SNVs. Following annotation with SpliceAI, only 21 were seen to have a SpliceAI score of 0.5 or greater (Figure 1A). Incidence of this class of variant in individuals recruited with FTAAD (9/703; 1.3%) was significantly elevated compared with that seen in non-FTAAD participants (12/77,492; 0.015%) and based on Fisher’s exact test, this enrichment was highly significant (OR=84, *p*=9.7×10^−14^). Although 0.5 is the intermediate cutoff recommended for SpliceAI,^20^ we explored the effects of varying this threshold for variant inclusion. Whilst the highest proportion of cases recruited under FTAAD was seen at the 0.8 cutoff (7/10, OR=260, *p*=5.4×10^−13^), the most significant *p-*value (11/34, OR=53, *p*=6.8×10^−15^) was observed at the 0.3 cutoff (Figure 1B).

**Figure 1:**
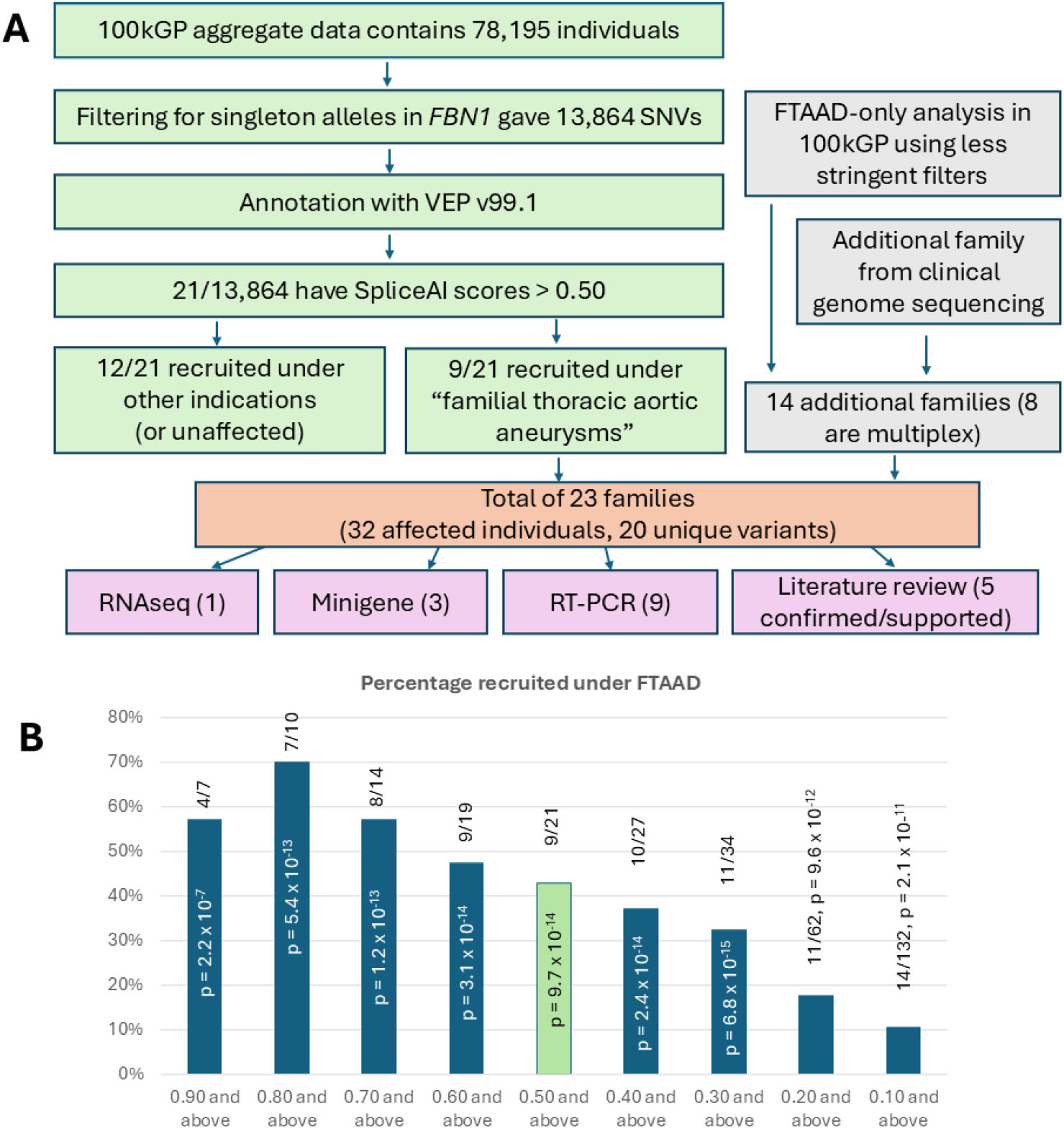
Analytical pipeline and phenotypic enrichment of rare splicing variants in *FBN1*. A) Green boxes denote steps taken in the initial analysis of data from the 100k Genomes Project (100kGP). Singleton SNVs were annotated with SpliceAI and filtered at 0.50 (AG, AL, DG, DL) as that is the recommended intermediate stringency cutoff. Grey boxes show subsequent steps taken to identify 14 additional families. The salmon box summarises the combined results from both arms of the study, whilst lilac boxes outline the different methods used to validate RNA effect and the numbers supported by each method. B) Chart showing percentage of rare splice carriers who were recruited under FTAAD at each level of SpliceAI cutoff. As the splice cutoff decreases additional FTAAD families were identified. The green box highlights the initial cutoff used as shown in panel A. The highest proportion of *FBN1* variant carriers being from the FTAAD group was seen at a 0.80 cutoff, but this would have resulted in Families 2, 7, 17 and 20 being overlooked. P values corresponding to Fisher’s exact test results of FTAAD enrichment for *FBN1* variant carriers show that the most significant enrichment was seen at the 0.3 cutoff.

The 703 individuals with FTAAD included in the primary analysis were from a total of 605 families, of which 523 (86%) were recruited with just a single affected individual. However, there were also 70 families with two affected individuals, 10 affected trios, 1 affected quad and 1 affected sextet. Therefore, we performed a secondary analysis including all families recruited to the 100kGP with FTAAD. In this analysis, we no longer required variants to be unique in the dataset and reduced the SpliceAI threshold to 0.2. We also included families that had previously been available only on GRCh37 (at the time of data aggregation) and one family sequenced as part of the GMS programme. This FTAAD-only analysis led to the identification of 11 non-canonical *FBN1* splice variants in 23 more affected individuals from 14 unrelated families.

In combination with the initial 9 variants, a total of 20 unique variants were identified in 32 individuals from 23 families (Table 1). These variants were spread throughout the gene from intron 1 to intron 63 (Figure 2A). Although there was no significant clustering, introns 12, 54 and 56 each harboured two variants. Overall, 14/20 variants lay beyond the ±8 splice regions used for prioritisation in the primary analysis performed by Genomics England and for 9 of these, inclusion of a pseudoexon (PE) was the predicted consequence. Exon extension was also the predicted effect for 9/20 variants, in one case involving a 5’-UTR exon (Figure 2B). Predictions of exon skipping and exon contraction were each observed just once. No coding variants predicted to result in anomalous splicing were identified.

**Table 1:**
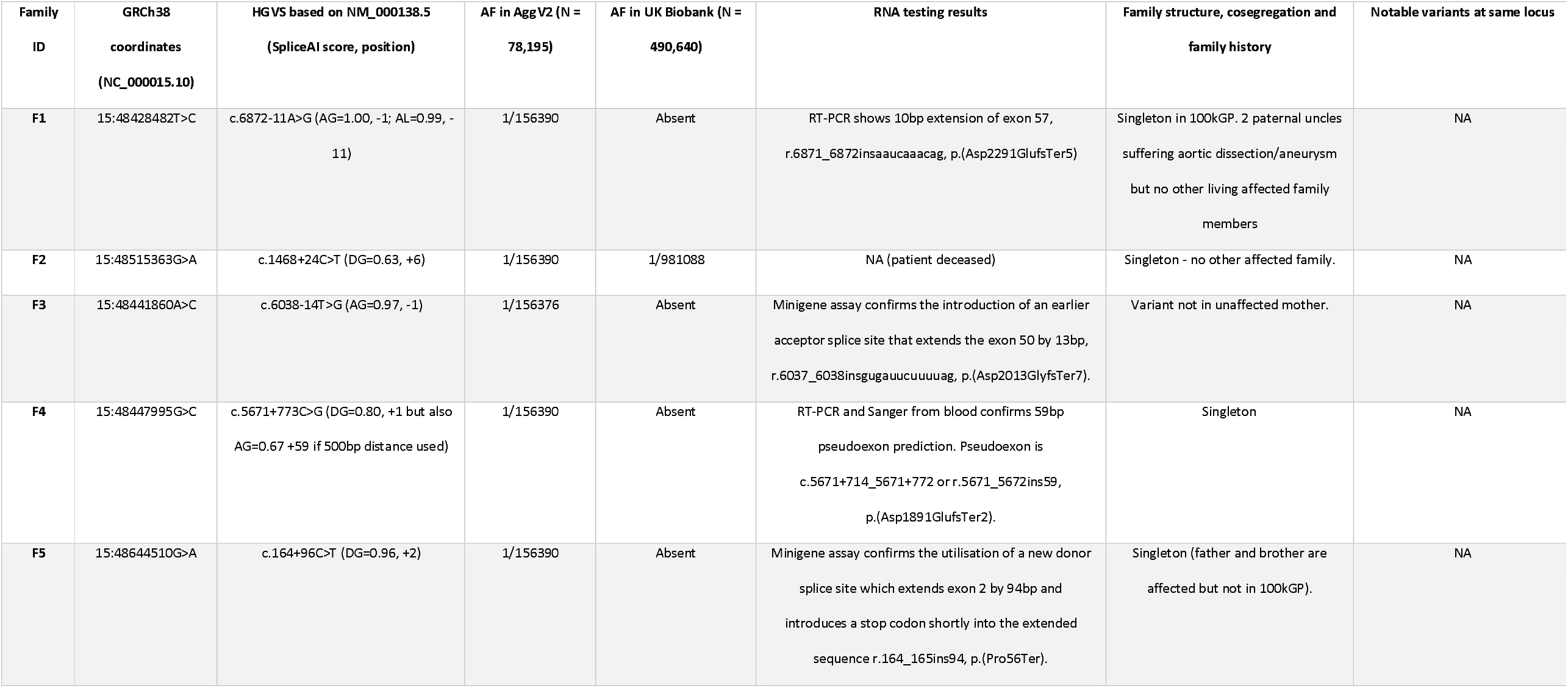
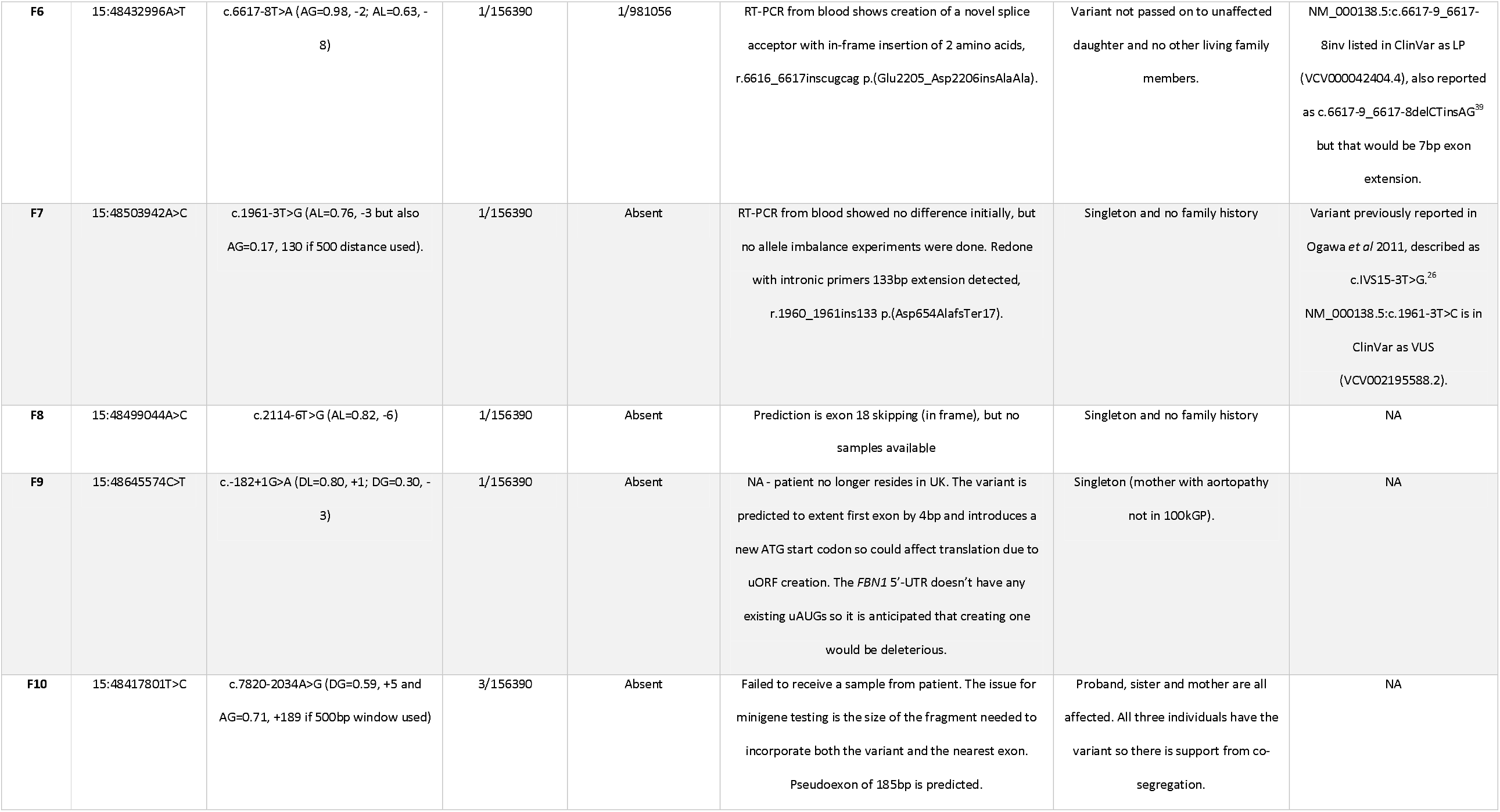
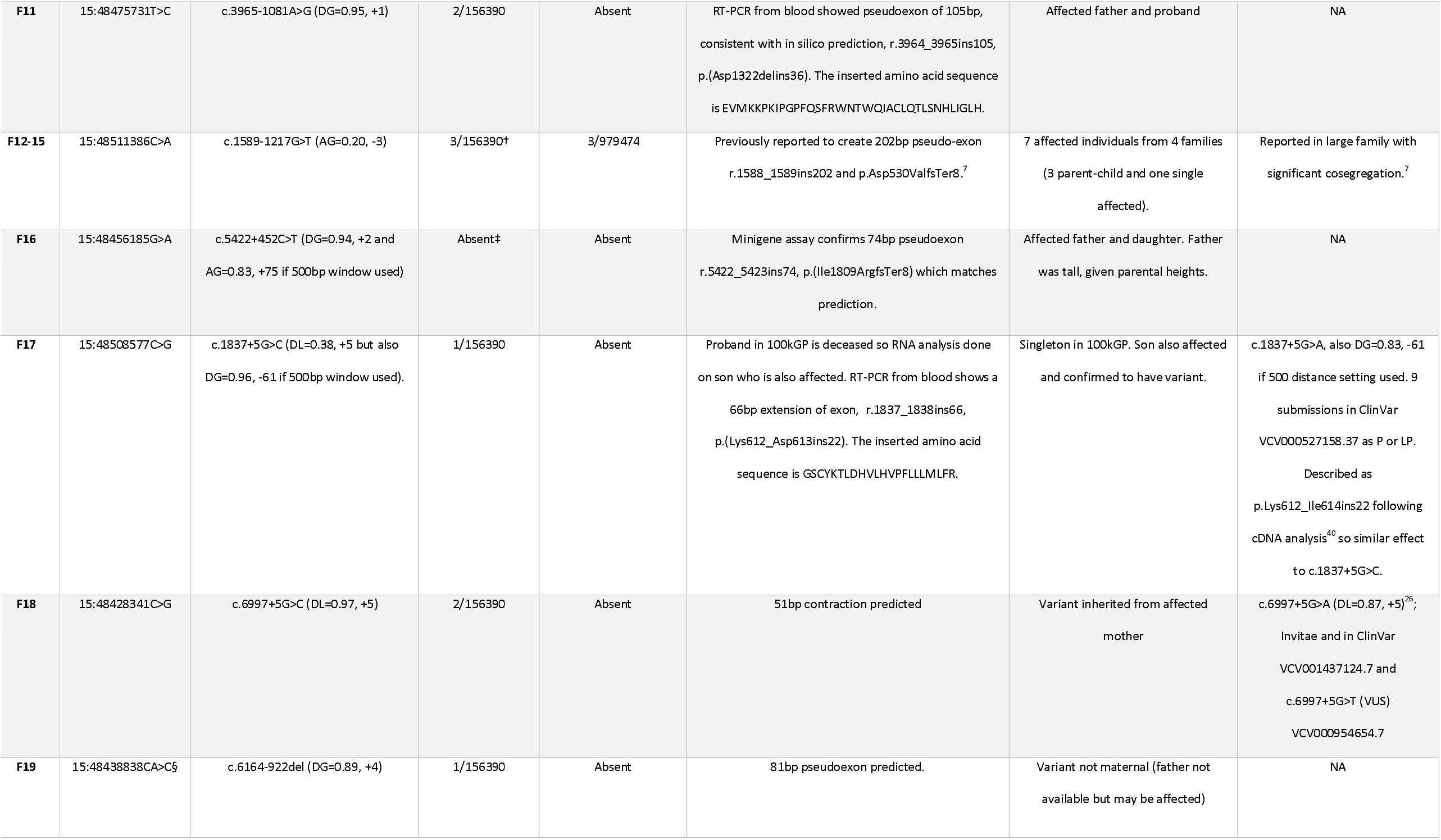
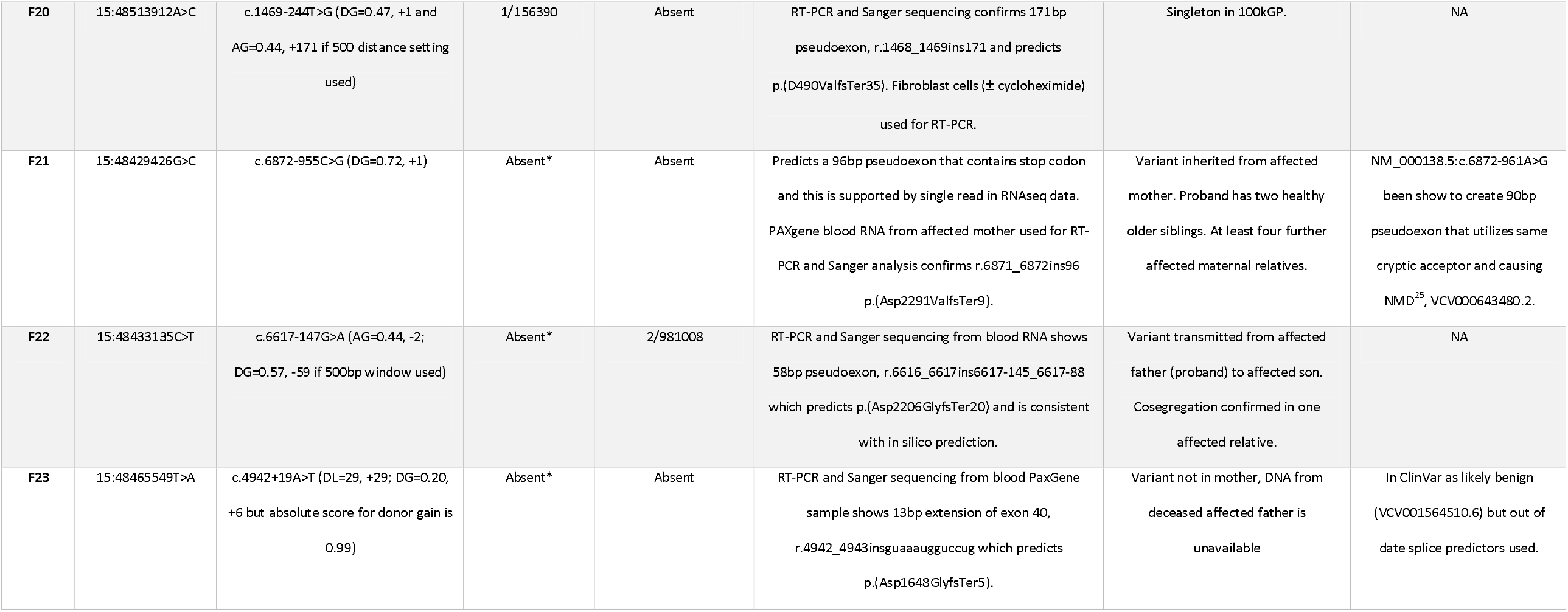
Summary of *FBN1* splicing *v*ariants in 23 families recruited with Familial Thoracic Aortic Aneurysm identified from the 100k Genomes Project or the Genome Medicine Service. AF, allele frequency. †only 3/7 individuals available on GRCh38 when AggV2 file produced. ‡Participants from the Genome Medicine Service programme are not included in AggV2. *data analysed on build GRCh37 so not in AggV2. § can also be described as NC_000015.10:g.48438840del.

**Figure 2:**
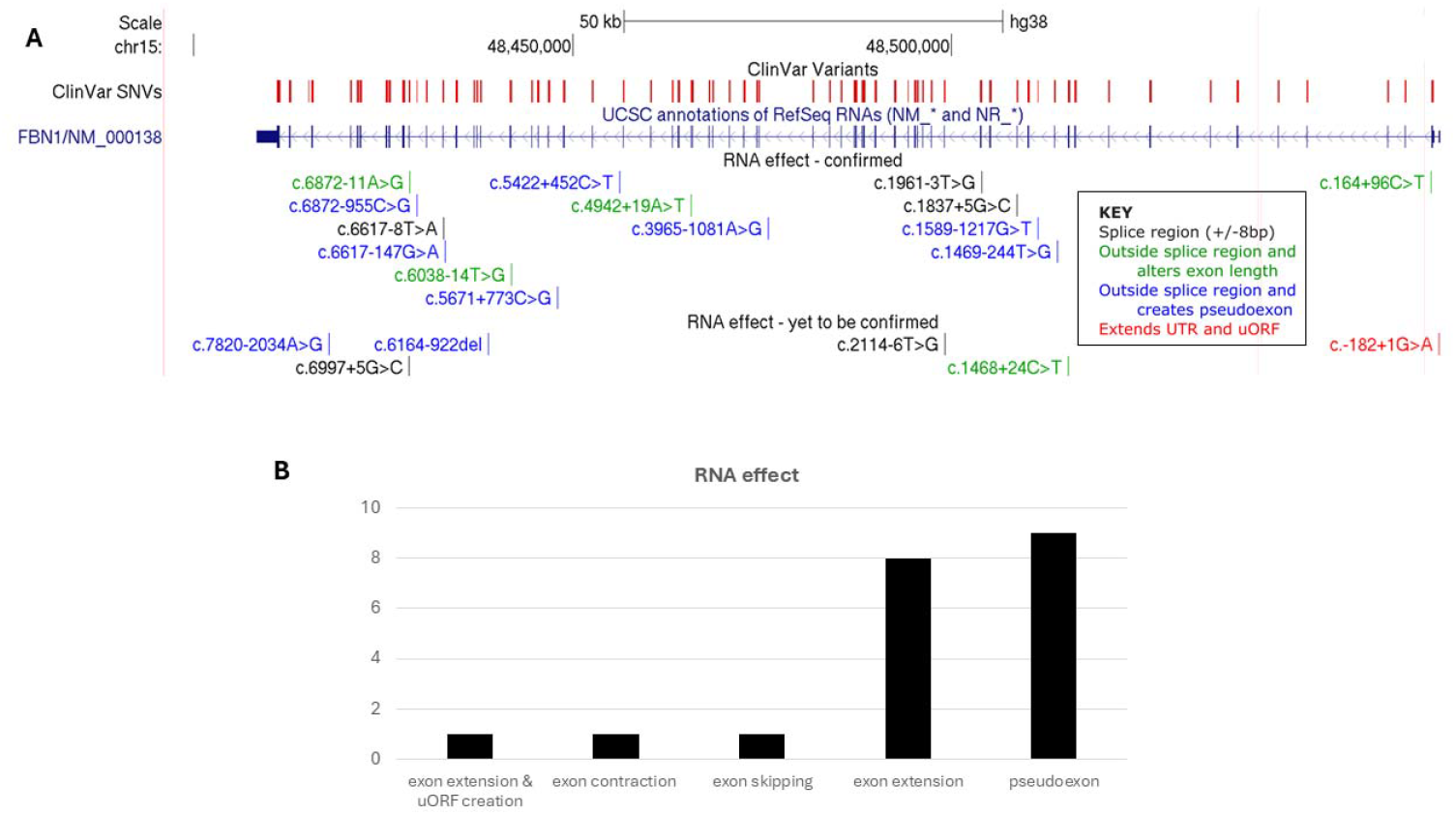
Distribution of noncanonical *FBN1* splicing variants. A) UCSC genome browser graphic showing that variants identified were spread throughout the gene. For viewing purposes, the multiregion view option (15:48396313-48540476, 15:48593651-48615606 and 15:48643624-48648334; GRCh38) was used to artificially shorten introns 2 and 6. An interactive UCSC custom session is available at https://genome.ucsc.edu/s/AlistairP/FBN1_distribution. The ClinVar track highlights that the vast majority of P/LP variants lie in or close to exons, with the only deep intronic variants shown are c.8051+375G>T, c.6872- 961A>G and c.1589-1217G>T. Of the variants described here, only c.1589-1217G>T and c.1961-3T>G were reported previously.^7,26^ B) Bar chart summarising the numbers of variant falling into each category of splice effect, with the most common consequence being creation of a pseudoexon and exon extension. HGVS annotations are based on transcript NM_000138.5.

The population allele frequencies for this set of variants in gnomAD v4 ranged from 0 up to 7/1,613,362 for the NM_000138.5:c.1468+24C>T variant in Family 2 (Table 1). The c.1468+24C>T variant is also one of six where RNA testing was not possible and so this remains a variant of unknown significance (VUS). Delta SpliceAI scores ranged from 0.20 in Families 12-15 (NM_000138.5:c.1589-1217G>T) and Family 23 (NM_000138.5:c.4942+19A>T) up to 1.00, predicting a change in acceptor site usage for NM_000138.5:c.6872- 11A>G in Family 1 (Figure S1). An interactive UCSC session showing the position of all 20 variants, SpliceAI-visual tracks showing absolute scores in comparison to the reference and additional tracks showing the predicted consequences of each variant, is available at https://genome.ucsc.edu/s/AlistairP/FBN1_pseudoexons.

### RNA testing for confirmation of aberrant splicing

Short read RNAseq using blood had already been performed for 10/23 families with candidate splice variants in *FBN1*. Coverage at the *FBN1* locus was extremely sparse (Figure S2). However, closer scrutiny of the loci of interest in the ten datasets identified one read in one individual that supported the predicted splice event, with no coverage for the relevant splice junction in the other 9 datasets. The only supported event was in the proband from Family 21, with a single read spanning NM_000138.5:c.6872-955C>G and the 3’ end of the predicted 96bp PE in intron 56 (Figure S3). The read alignment supported one of the two predicted splice junctions, with the soft clipped bases matching the sequence from the start of exon 57. Similar junctions were not observed in >400 control RNAseq datasets sequenced using the same pipeline (Figure 3A). The variant appears to activate the same cryptic acceptor site as described in a previous study^25^ (Figure 3B) and the PE was confirmed by RT-PCR using an independent blood sample (Figure 3C).

**Figure 3:**
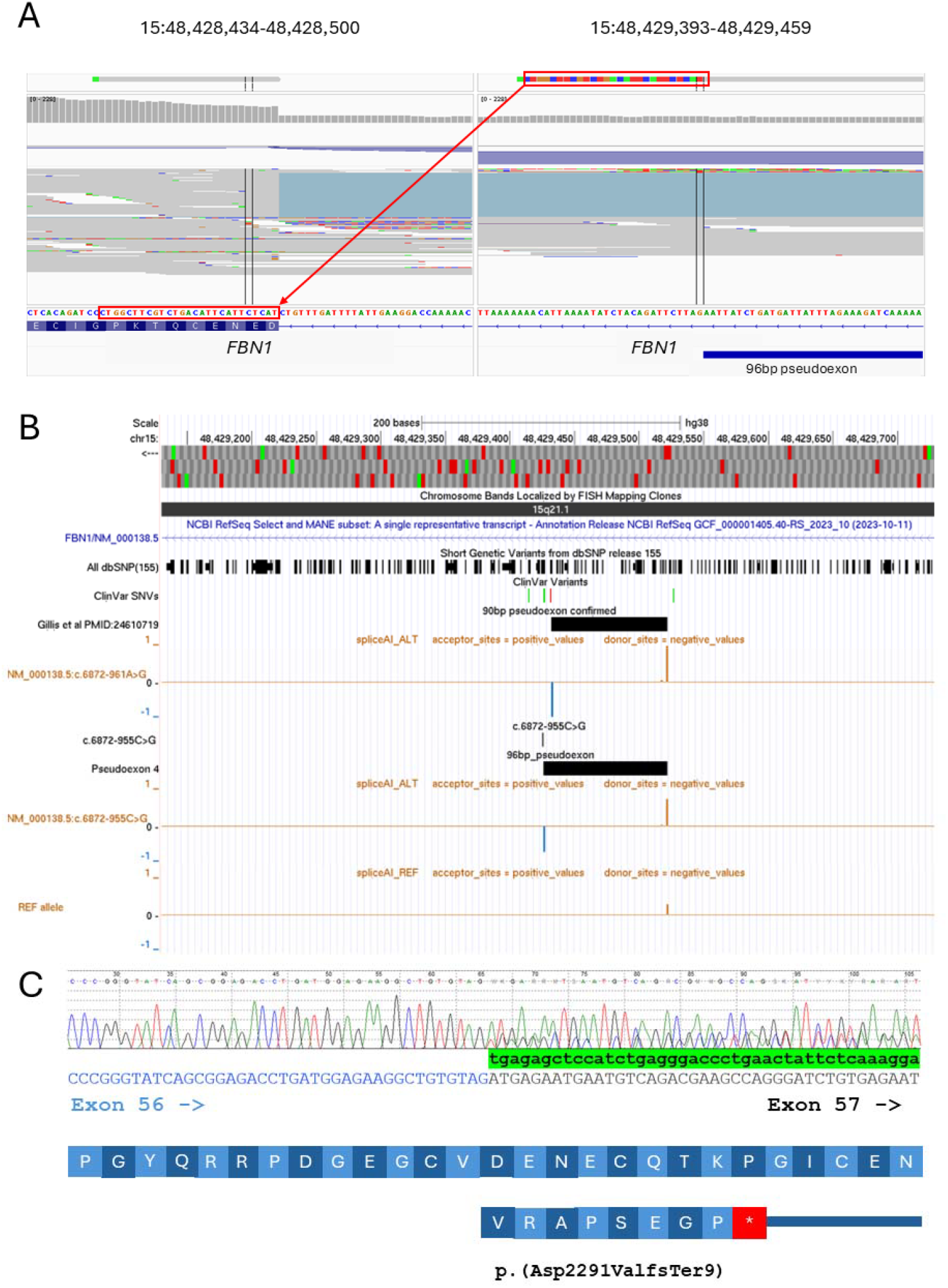
Deep intronic variant in Family 21 leads to activation of a dormant splice acceptor site and a 96bp pseudoexon. A) IGV screenshot showing single RNAseq read at the informative locus in intron 56. The red boxes highlight soft-clipped sequence which match the start of exon 57 and consistent with the SpliceAI prediction. The lower track shows squished view of reads from 402 merged RNAseq datasets run using a similar pipeline during the same calendar month. No similar junctions are observed. B) UCSC browser image showing positions of c.6872-955C>G and the absolute SpliceAI scores in comparison to the reference, as generated by SpliceAI-visual. The predicted pseudoexon activates the same dormant splice acceptor site that was upregulated by the c.6872-961A>G variant (red) shown in the ClinVar variant track. The positions of the two overlapping 96bp and 90bp pseudoexons are shown by black boxes. The dormant splice acceptor site has an absolute score of 0.29. C) PCR-Sanger sequencing of cDNA further supports the *in silico* prediction, with the sequence highlighted in green corresponding to the 5’ end of the 96bp pseudoexon. The inserted sequence is predicted to lead to a frameshift and termination after the insertion of 8 novel amino-acids.

For 9 families, RT-PCR was used as a standalone test for assessing the effect of the variant on the *FBN1* transcript (Figure 1A, Table 1). For 8/9 families, blood-derived RNA was used as a template and despite low expression in this tissue, informative results were obtained in every case. For Family 7, multiple rounds of primer design were required, but otherwise this approach proved to be highly efficient. RT-PCR requires fresh patient-derived RNA samples and it was not possible to obtain these for all participants. Therefore, to test additional variants and explore the feasibility for alternative approaches, minigene assays were considered. Minigene assays were performed for three families and in all cases confirmed the predicted splice events. These included variants in Families 3 and 5 resulting in exon extension of 13bp and 94bp respectively with both predicting the introduction of PTCs (Figure 4A,B). For Family 3, blood samples later became available but instead of confirming the 13bp extension, RT-PCR results suggested the occurrence of NMD (Figure S4). Minigene testing for the heterozygous NM_000138.5:c.5422+452C>T variant found in Family 16 confirmed the prediction of a 74bp PE (Figure S5) and the insertion, r.5422_5423ins74, would create a PTC, p.(Ile1809ArgfsTer8).

**Figure 4:**
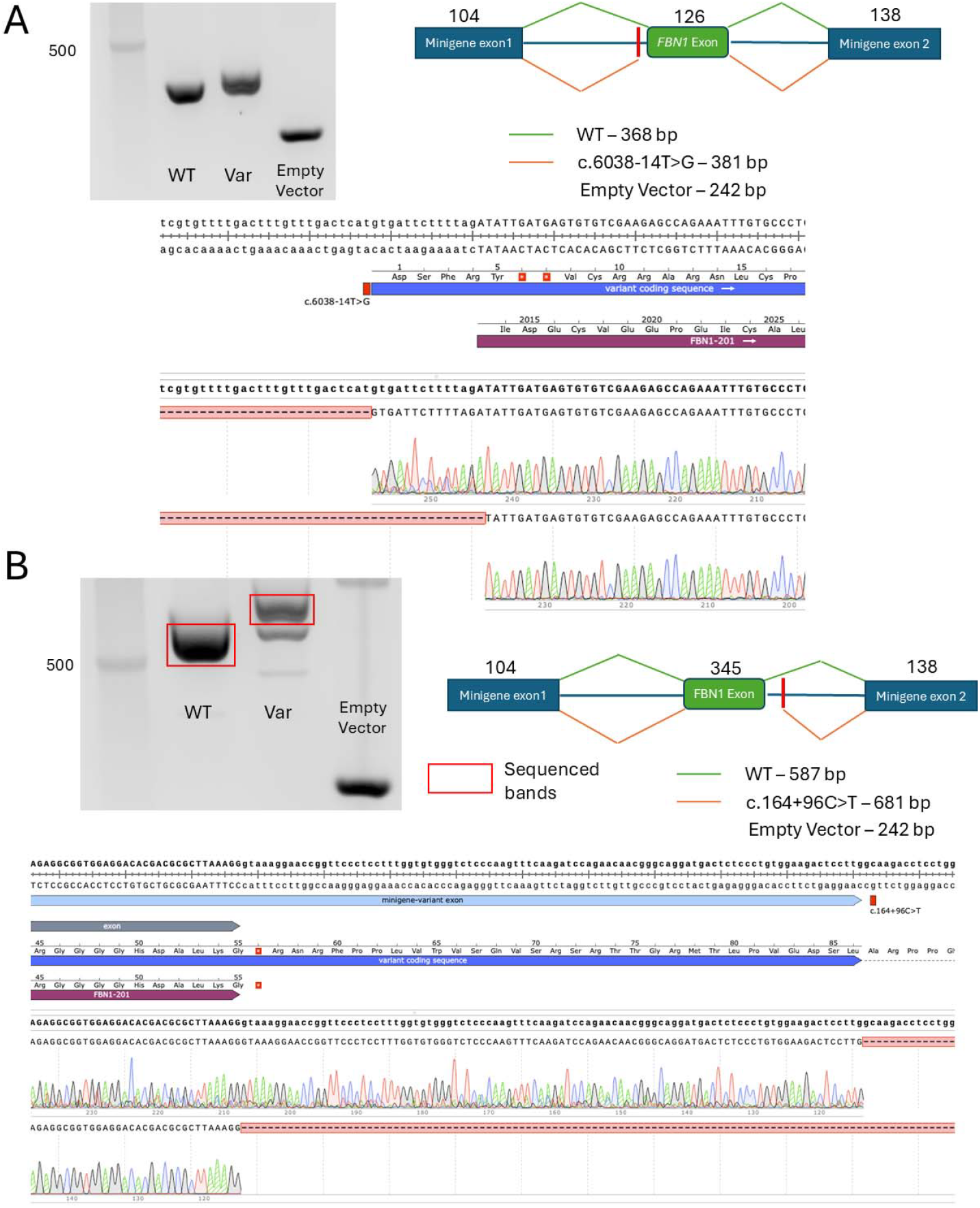
Minigene assay results confirm *in silico* predictions of exon extension for Families 3 and 5. A) Agarose gel electrophoresis showing aberrant RT-PCR product associated with the c.6038-14T>G variant in Family 3 due to the introduction of an earlier acceptor splice site consistent with the SpliceAI prediction for a novel acceptor site (AG=0.97, −1 and AL=0.20, −14). This extends exon 50 by 13bp, r.6037_6038insgugauucuuuuag and predicts a frameshift p.(Asp2013GlyfsTer7). B) Gel image and Sanger sequence data showing that the c.164+96C>T variant in Family 5 results in the utilisation of a new donor site which extends exon 2 by 94bp and is consistent with the SpliceAI prediction of the introduction of a novel deep intronic donor splice site (DG=0.96, +2 and DL=0.45, +96). This introduces a stop codon shortly into the extended sequence, r.164_165ins94, p.(Pro56*).

Overall, experimental confirmation of the putative splice variants has now been performed for 14 of 20 variants, 13 as part of the present study and one variant (Families 12-15) where RNAseq data had already been reported in the literature.^7^ In all cases, aberrant splicing was detected. Overall, we estimated that minigene assays required approximately twice the resources in terms of costs and hands-on time, compared with the simpler RT-PCR approach (Supplementary Methods).

### Interpretation of non-canonical splice variants based on *in silico* approaches

The ability to detect variants likely to impact splicing is essential, but accurate prediction of the alternative splice event is highly beneficial for variant interpretation and for designing appropriate confirmatory RNA testing assays. Typically, the SpliceAI scores used are the delta scores, representing the difference score between reference and variant alleles.^19^ In this analysis, we observed the importance of more refined interpretation of candidate splice variants.

Although precalculated SpliceAI scores using the 50bp window are the scores typically used by researchers, and used in our initial prioritisation steps here (Figure 1A), for further interpretation the window size was expanded to 500bp. Strikingly, for three families, the more distant SpliceAI prediction increased the maximum delta SpliceAI score (Table 1). The clearest example of an increased SpliceAI score using a larger analysis window is for Family 17. The proband and her affected son harboured the NM_000138.5:c.1837+5G>C variant which has a moderate score for a donor loss (DL=0.38, +5), but the score increases to DG=0.96 (−61) if the distance setting is increased. RT-PCR using blood-derived RNA from the son confirmed an in-frame 66bp extension of exon 15, r.1837_1838ins66, p.(Lys612_Asp613ins22) (Figure S6). Similarly, in Family 22, NM_000138.5:c.6617-147G>A has a moderate prediction for an acceptor gain (AG=0.44, −2) but with the larger window, there is a stronger prediction for a donor gain (DG=0.57, −59) and together this would result in a PE of 58bp. RT-PCR and Sanger sequencing from blood-derived RNA confirmed this PE, r.6616_6617ins6617- 145_6617-88, p.(Asp2206GlyfsTer20) (Figure S7). A third example of this occurrence was for Family 10 where *in silico* analysis with the larger analysis window predicted a 185bp PE (Table 1).

In one case, a more accurate splicing prediction was able to overcome some of the difficulty with confirmatory RNA testing, even though the maximum SpliceAI score was unchanged. The NM_000138.5:c.1961-3T>G variant identified was described in an independent study but RNA testing was not undertaken.^26^ For Family 7, prior genetic testing using a clinical exome had detected the same variant. However, RNA testing was not indicative of aberrant splicing, but it had been noted that a limitation of the RT-PCR experiment was that the possibility of a large insertion and PCR-bias or transcript degradation due to NMD could not be excluded. Thus, the variant had remained a VUS. Closer scrutiny of the SpliceAI scores using a 500bp analysis window showed that whilst the main effect was likely loss of an acceptor site (AL=0.76, −3), there was also a weaker cryptic acceptor site predicted 130bp away (AG=0.17, 130). Although the delta SpliceAI score was <0.2, the raw reference allele has a 0.06 score at this position and so the absolute SpliceAI score for the variant was 0.23. Repeating the RT- PCR assay using customised primers confirmed a 133bp extension of exon 17 in the 5’ direction, r.1960_1961ins133, consistent with the prediction (Figure S8). This predicted a frameshift leading to a PTC, p.(Asp654AlafsTer17).

Using the delta scores alone can also limit variant interpretation^19^, especially when there is potentially a high propensity for splicing with the reference allele (and therefore a high raw score). The clearest example of the importance of assessing absolute SpliceAI scores is seen in Family 23. In this Family, the c.4942+19A>T variant which is listed as likely benign in ClinVar (VCV001564510.6) was detected in the proband. Although the delta score is low (DG=0.20, +6), the raw SpliceAI splice-donor scores increase from 0.79 to 0.99 with the variant (Figure S9A), resulting in a higher score than the canonical donor site which decreases from 0.99 to 0.84. If assessing just the delta score, this variant would be easy to overlook. RT-PCR and Sanger sequencing confirmed a 13bp extension of exon 40, r.4942_4943insguaaaugguccug which predicts p.(Asp1648GlyfsTer5) (Figure S9B).

### Examples of variants identified

We observed 13 putative loss of function alleles resulting from both inappropriate extension of canonical exons and through the creation of novel PEs. For 7/20 variants, a splicing anomaly was observed/predicted that is not expected to result in a PTC. These variants include in-frame indels (2 deletions and 4 insertions, Table S4), and one variant expected to impact the 5’-UTR. Subsequent interpretation of these variants requires in depth knowledge of the protein structure and functional domains, and/or further functional assessment. In Family 6, the c.6617-8T>A variant was shown to extend exon 55 by 6bp and this predicts the insertion of two alanine resides, the structural consequences of which are hard to determine. In Family 8, in-frame exon skipping is predicted, whilst in Families 11 and 19 the PEs predict in-frame insertions of 35 and 27 novel amino-acids, respectively. The 66bp exon extension in Family 17 and 51bp contraction in Family 18 are also in-frame.

Another notable example is the NM_000138.5:c.-182+1G>A variant detected in Family 9. This variant involves a non-coding exon in the 5’-UTR, but is strongly predicted to alter splicing of this exon (DL=0.80, +1; DG=0.30, − 3), potentially extending exon 1 by 4bp. If correct, this same variant would then also create an AUG codon and an upstream Open Reading Frame (uORF) that would encode a short peptide of 27 amino-acids. The 5’-UTR of *FBN1* does not contain any other uORFs and so we hypothesize that this variant could alter the efficiency of translational initiation from the canonical start-site. The role of uORFs in translational regulation of protein expression among humans is becoming increasingly well understood^27^, but confirmation of such a mechanism would require further functional analysis, such as with luciferase reporter assays. This individual had a MFS phenotype, including non-progressive dilated aortic root (4.0cm), pectus and arachnodactyly and his mother is similarly affected. Previous testing of *FBN1* by Sanger sequencing and MLPA had returned negative results, but unfortunately segregation/RNA testing in the family has not been possible. We are not aware of any similar examples of SNVs that simultaneously alter both transcription and translation in the literature.

Of the 20 variants identified, only one (NM_000138.5:c.1589-1217G>T) was recurrent and identified in 7 individuals from 4 independent families (Families 12-15), each recruited from a different geographical region of the UK. Despite the relatively low SpliceAI score (AG=0.20; rising to DG=0.32 if absolute scores and larger analysis window is used), the variant was prioritised on account of the 7 individuals harbouring this variant being recruited under FTAAD and the variant not being present in any other individuals recruited to the 100kGP. In parallel with this observation, c.1589-1217G>T was detected by Guo *et al* following linkage analysis performed in a large MFS pedigree and was shown to result in a novel 202bp PE by RNAseq.^7^ The variant is also listed in ClinVar under accession VCV002664316.1. Since this variant has now been identified in at least five families, we performed haplotype studies as described previously^21^, to assess whether the variant lay in a shared segment that is identical-by-descent. This included comparison of data for the 100kGP families to a set of 16 rare SNVs found on the disease haplotype in the original TAA758 family (pers. comm. Dong-Chuan Guo).^7^ Altogether, we found no strong evidence supporting a single founder origin. Furthermore, we identified a nearby SNP (256bp away) that could be phased with read-level data. In Families 12, 14 and 15 c.1589-1217G>T was *in cis* with the reference allele at rs9806163, whereas in Family 13 it was *in cis* with the non-reference allele (Figure S10). Further details of haplotype studies performed and gnomAD findings are mentioned in a Supplementary Note. Together, these data suggest independent mutational recurrence for c.1589-1217G>T. It is therefore important to screen for this cryptic variant in MFS cohorts from all ethnicities.

### Clinical impact of non-coding variants in FBN1

We estimate the diagnostic yield based on the set of non-canonical *FBN1* splicing variants described here in the FTAAD cohort from the 100kGP to be 2.8% in the primary analysis (i.e. 17 unrelated probands from 605 FTAAD families included in AggV2) or 3.3% (22/672, based on latest 100kGP release) in the FTAAD-only analysis. In all families where data are available, *FBN1* testing had been undertaken prior to recruitment to the 100kGP. Many families had undergone a long diagnostic odyssey as evidenced by the wide range of methods used for prior testing that included DHPLC, PCR-Sanger, NGS panel testing, MLPA and clinical exome analysis. Aortic root dilatation ranged from being in the normal range for several individuals, up to 9cm in diameter (prior to surgery) for the proband in Family 3. Cardiac surgery had been performed in at least 11/23 families. Chest abnormalities were described in 15/23 families whilst *ectopia lentis* was reported only in 5/23. Striae were noted in 8/23 and standing heights of 190cm and above were reported for 9/23 families. Further clinical details are available in Table S4.

Whilst the genome sequencing used in this study revealed the likely genetic diagnosis for a total of 32 individuals from 23 families, it is anticipated that cascade testing will be ongoing for several years and will identify many other at-risk individuals who may benefit from regular cardiac surveillance. The best example is Family 22, where the pedigree stretches over four generations and there are 10-20 individuals for whom cascade testing for the c.6617-147G>A variant is anticipated (Figure S7C). Similarly for Family 21, although the full pedigree was not available for review, this nuclear family is part of another large MFS kindred, with at least four further affected maternal relatives reported, hence ongoing cascade testing for the c.6872-955C>G variant will impact directly on cardiac surveillance.

### Replication using UK Biobank

Just under 1% (5,000/502,284) of UKB participants had the ICD10 code I71 “Aortic aneurysm and dissection” listed either in their Hospital Episode Statistics (HES; n=4736) or in the death record (n=264). The incidence of aortic aneurysm in the general population is thought to be 1-2% above the age of 65^28^ and so broadly in line with data from the UKB. There were also 112/502,284 participants with the code of Q87.4 (MFS) listed in the HES data. The overlap of these codes was surprisingly low, with only 40/112 of the individuals with Q87.4 also having the I71 code. There were 84 ultra-rare SNVs identified with SpliceAI delta scores of 0.50 or more (Table S5) found in a total of 146 UKB participants (145 heterozygotes and 1 homozygote). Rare *FBN1* splice variants were not significantly enriched in the I71 cohort (OR=1.38, *p*=0.66), whereas for the more specific Q87.4 code there was a highly significant enrichment (OR=131, *p*=4.15×10^−8^) and one of the four contributing variants was a variant NM_000138.5:c.863-3641A>G (DG=0.79, 1) in intron 8 and predicted to create a 485bp PE. The individual harbouring this deep intronic variant did not carry any other likely-pathogenic variants in *FBN1*. Due to the nature of UKB consent, we were unable to obtain any further details or fresh biological samples to perform RNA confirmatory studies.

## DISCUSSION

Several case reports have indicated that intronic variants in *FBN1* can result in aberrant splicing and provide a molecular diagnosis for families with MFS.^7,25,29,30^ However, the overall contribution of such variants to the etiology of MFS is unclear. In this study, the availability of genome sequencing data across a large clinical cohort that included a large number of participants with a diagnosis of FTAAD allowed us to explore the incidence of splice variants in genetically unsolved cases. Amongst >600 FTAAD families in the 100kGP without a previously identified molecular diagnosis, ~3% were found to have a candidate intronic small variant and for 9/20 variants, the creation of a PE was predicted. Extrapolation of this yield into other clinical cohorts is difficult. Firstly, MFS was not a specific diagnostic category used for recruitment to the 100kGP and so it is difficult to estimate how many of the 672 families recruited with FTAAD also had a clinical suspicion specifically for MFS. Secondly, prior genetic testing was a requirement for recruitment to the 100kGP and since many families were recruited through clinical genetics, it is possible that this collection of unsolved families is biased towards those with a strong family history and/or clinical suspicion of a monogenic disorder. As a result, non-canonical splice variants are likely better represented here than would be the case in MFS cohorts (or unselected population groups such as UKB) naïve to previous genetic testing. We also note that since a single splice prediction tool was used for variant prioritisation, there could be additional splice variants in *FBN1* among the families with FTAAD in the 100kGP that were not considered. The fact that no exonic splice variants were identified may be the result of poorer predictive capabilities for splice variants in exonic regions.^31^

We observed a strong enrichment (OR=84, *p*=9.7×10^−14^) of singleton splice SNVs in the FTAAD cohort of the 100kGP. However, we failed to replicate this observation among individuals with “Aortic aneurysm and dissection” in the UK BioBank. This lack of replication may be due to recruitment bias in the 100kGP, or a lack of phenotypic specificity in the UK BioBank cohort, in which the ICD10 code, “Aortic aneurysm and dissection” is less precise than the clinical indication in the 100kGP dataset (where families were recruited with FTAAD, i.e. “Familial Thoracic”). Abdominal aortic aneurysms are more common and usually related to age, male sex and atherosclerosis and are thought not to have as significant a genetic contribution. The aortic aneurysm and dissection ICD10 code we used does not differentiate between the thoracic and abdominal aorta. UKB also contained 112 individuals with the more explicit ICD10 code of Q87.4 and for this phenotype, the enrichment of *FBN1* splice variants was highly significant (OR=131, *p*=4.15×10^−8^). Unfortunately, it is unclear whether Q87.4 is typically assigned a clinical or genetic basis and if the latter then this supporting evidence is circular and should be treated with caution.

For 14/20 of the candidate variants identified, an impact on mRNA splicing has now been confirmed using at least one RNA testing method. A limitation remains in this study that for the remaining six variants, RNA testing was not possible for a variety of reasons, including patients not being available to provide samples for testing. However, this limitation highlights challenges with confirmatory RNA testing, especially for genes with low expression in blood, for which bespoke testing methods or invasive sample collection approaches may be required. In this study, we made use of a number of different approaches for RNA analysis and each method comes with its own set of advantages and disadvantages. RT-PCR using RNA extracted from blood was the approach that requires the least resources and was the most commonly used approach used in this study. This technique was performed for eight families as a standalone test and in two further families for confirmation of minigene/RNAseq results. RT-PCR using RNA derived from patient fibroblasts was used in a further case and this approach allows the introduction of NMD inhibitors such as cycloheximide to the culture media to help monitor the impact of NMD and potentially enhance any signal from aberrant transcripts. Although fibroblasts have traditionally been used for RNA analysis of *FBN1*, a recent study proposed urine analysis as a viable non-invasive solution.^32^ Minigene assays require more resources but are helpful for patients for whom collection of fresh samples is not feasible. However, effective design of minigene constructs can be problematic for variants that lie too far into an intron and testing splice donor variants involving exon 1 are not straightforward to design as these exons do not harbour natural splice acceptor sites. Although these can be engineered^33^, this procedure is not trivial. Based on the results presented here, RNAseq from blood is not recommended for analysis of splicing variants in *FBN1* and the single read in data from Family 21 capturing one of the predicted PE junctions (Figure 3A) was fortuitous.

Pathogenic variants involving *FBN1* are highly actionable in terms of surveillance/management and the gene is included in the ACMG list recommended for reporting of secondary findings.^34^ It is therefore critically important that RNA validation studies are performed for confirmation of pathogenicity. Currently, provision of follow up RNA testing is variable in existing clinical testing pathways and so many families had experienced long diagnostic odysseys. For two families presented here, the variant of interest was within 15bp of the nearest exon and had been previously identified using a clinical panel test. However, in the absence of confirmatory RNA analysis, the variants were considered as VUS, and for one family additional genetic testing was pursued. Increased availability of RNA testing would help resolve variants of uncertain significance and in some cases, negate repeated DNA testing. Widespread use of RNA testing is hindered by low *FBN1* expression in blood where the median transcript per million value is just 0.2158 (803 datasets from GTEx; https://gtexportal.org/home/gene/FBN1). In this study, most RNA experiments were performed using RT-PCR by a single clinically accredited genomics laboratory that has significant expertise in working with low abundance transcripts.

Over the last decade there have been vast improvements in the accuracy of splice prediction algorithms and SpliceAI, which uses a deep neural network to facilitate highly accurate prediction of splice junctions from primary sequence^20^, is one of the most widely used approaches. Our results demonstrate that it is now feasible to scan large intronic regions across thousands of individuals in the search for cryptic splice variants. For the set of variants presented here, *in silico* predictions all proved to be correct. In contrast, a study from 2012 performed RNA analysis for 36 *FBN1* variants and identified only two that caused an abnormality of splicing, leading to the conclusion that, at that time *in silico* analysis did not accurately predict the splicing seen experimentally.^35^ Further improvements in splice prediction tools may enable the strength of evidence attributed to computational evidence and variant co-localisation with other previously reported variants to be increased and potentially reduce the need for complex RNA testing. An example is Family 18; although RNA studies were unable to be performed, support for pathogenicity was obtained from variant colocalization at the same +5 position. The proband and affected mother both harboured a NM_000138.5:c.6997+5G>C variant that predicted a strong donor loss (DL=0.97, +5) and a donor gain (DG=0.57, +56) that would result in contraction of the exon by 51bp. Database searches indicated that another variant at the same base c.6997+5G>A with strikingly similar *in silico* prediction (DL=0.87, +5, DG=0.58, +56) has been described previously^26^. The variant is also listed in ClinVar with one likely-pathogenic assessment (VCV001437124.7). For some genes, deep intronic splice variants can show significant clustering due to the intronic sequence already having exon-like properties and thus only requiring a small increase in splice efficiency to result in PE inclusion.^36,37^ Such information should be incorporated into the analysis of candidate splice variants.

However, our experience demonstrates that adapting the use of SpliceAI predictions from the default parameters facilitates a more refined analysis of candidate splice variants that increases diagnostic yield and enables improved experimental design of confirmatory RNA analysis. In particular, we note the benefits of increasing the SpliceAI window size from 50bp to 500bp and an analytical window of even 5,000bp has been proposed elsewhere.^38^ Here, the increased window size was important for determining the formation of PEs as all nine PEs detected were over 50bp in size (58bp to 202bp, Table 1). However, when designing analytical pipelines it is important to balance gains in analytical accuracy with the bioinformatic resources required and the interpretation burden. Highly customised modification of parameters is not feasible at scale in high throughput analysis pipelines, highlighting that further improvements in splice prediction tools would be beneficial.

In the case of *FBN1*, analysis of candidate splice variants can be facilitated by the phenotypic specificity of MFS as the associated condition. For other disorders with a broader phenotypic range and higher genetic heterogeneity, bespoke adaptation of splice parameters with the aim of achieving high sensitivity would be substantially more challenging, likely resulting in a greater analytical challenge and potentially more false positives.

In conclusion, our systematic evaluation of candidate splice variants in *FBN1* in the 100kGP highlights the benefits of considering rare non-coding variants, prioritised by computational predictions. Meticulous variant analysis led to *in silico* predictions proving to be correct in all cases where RNA testing was possible. Our findings indicate that ultra-rare *FBN1* variants with supportive SpliceAI scores should be considered as strong candidates in individuals with a FTAAD phenotype and/or suspicion for MFS, indicating that WGS would be an appropriate test method for families without a candidate diagnosis identified. However, for any increased yield to be realised, such changes must also be accompanied by an increased capacity for splicing analysis of low abundance genes being available in clinical laboratories.

## Data availability

Data from the 100kGP and the GMS is held in the National Genomic Research Library, Genomics England, https://doi.org/10.6084/m9.figshare.4530893.v7. Data from UK Biobank cannot be shared publicly because of data availability and data return policies. Data are available from the UK Biobank for researchers who meet the criteria for access to datasets to UK Biobank (www.ukbiobank.ac.uk).

## Acknowledgements

We thank all families from the 100kGP and GMS for their involvement in this study and the late Bruce Castle for clinical input. This research was made possible through access to data in the National Genomic Research Library, which is managed by Genomics England Limited (a wholly owned company of the Department of Health and Social Care). The National Genomic Research Library holds data provided by patients and collected by the NHS as part of their care and data collected as part of their participation in research. The National Genomic Research Library is funded by the National Institute for Health Research and NHS England. The Wellcome Trust, Cancer Research UK and the Medical Research Council have also funded research infrastructure. This research has been conducted using the UK Biobank Resource under Application Number 103356 and uses data provided by patients and collected by the NHS as part of their care and support. UK Biobank protocols were approved by the National Research Ethics Service Committee.

## Author contributions

ATP, IB, GH, CS and SW performed analysis of GS data; DJB, JH, HW, CK, YK and MD performed RT-PCR splicing assays and HBT performed minigene experiments. EB, SLC, CF, JE, RW, SFS, HC, PC, MM, MP, SA, DM-R, JD, PM, AD, AS, KP, LARK, RT, CT, SE, CS-S, JF, VC, SH, SS and CM provided clinical information and helped to recruit these families to the 100kGP. RTO’K, SB, DB, ARW MNW, NST, ELB and JCT reviewed data and provided oversight to study. ATP and SW wrote the manuscript which was reviewed by all authors.

## Ethics Declaration

Ethics approval was from Cambridge South REC (14/EE/1112).

## References

1. Marfan, A.B. (1896). Un cas de déformation congénitale des quatre membres, plus prononcée aux extrémités, caractérisée par l’allongement des os avec un certain degré d’amincissement. Bulletins et Mémoires 13 (3rd series), 220–226.

2. Cui, R.Z., Hodge, D.O., and Mohney, B.G. (2023). Incidence and de novo mutation rate of Marfan syndrome and risk of ectopia lentis. J AAPOS 27, 273 e271–273 e274. 10.1016/j.jaapos.2023.07.006.

3. Groth, K.A., Hove, H., Kyhl, K., Folkestad, L., Gaustadnes, M., Vejlstrup, N., Stochholm, K., Ostergaard, J.R., Andersen, N.H., and Gravholt, C.H. (2015). Prevalence, incidence, and age at diagnosis in Marfan Syndrome. Orphanet J Rare Dis 10, 153. 10.1186/s13023-015-0369-8.

4. Loeys, B.L., Dietz, H.C., Braverman, A.C., Callewaert, B.L., De Backer, J., Devereux, R.B., Hilhorst-Hofstee, Y., Jondeau, G., Faivre, L., Milewicz, D.M., et al. (2010). The revised Ghent nosology for the Marfan syndrome. J Med Genet 47, 476–485. 10.1136/jmg.2009.072785.

5. Dietz, H.C., Cutting, G.R., Pyeritz, R.E., Maslen, C.L., Sakai, L.Y., Corson, G.M., Puffenberger, E.G., Hamosh, A., Nanthakumar, E.J., Curristin, S.M., and et al. (1991). Marfan syndrome caused by a recurrent de novo missense mutation in the fibrillin gene. Nature 352, 337–339. 10.1038/352337a0.

6. Dietz, H.C., Pyeritz, R.E., Puffenberger, E.G., Kendzior, R.J., Jr., Corson, G.M., Maslen, C.L., Sakai, L.Y., Francomano, C.A., and Cutting, G.R. (1992). Marfan phenotype variability in a family segregating a missense mutation in the epidermal growth factor-like motif of the fibrillin gene. J Clin Invest 89, 1674–1680. 10.1172/JCI115766.

7. Guo, D.C., Duan, X., Mimnagh, K., Cecchi, A.C., Marin, I.C., Yu, Y., Velasco, W.V., Lee, K., Zhu, X., Murdock, D.R., et al. (2023). An FBN1 deep intronic variant is associated with pseudoexon formation and a variable Marfan phenotype in a five generation family. Clin Genet 103, 704–708. 10.1111/cge.14322.

8. Dietz, H.C., Saraiva, J.M., Pyeritz, R.E., Cutting, G.R., and Francomano, C.A. (1992). Clustering of fibrillin (FBN1) missense mutations in Marfan syndrome patients at cysteine residues in EGF-like domains. Hum Mutat 1, 366–374. 10.1002/humu.1380010504.

9. Muino-Mosquera, L., Steijns, F., Audenaert, T., Meerschaut, I., De Paepe, A., Steyaert, W., Symoens, S., Coucke, P., Callewaert, B., Renard, M., and De Backer, J. (2018). Tailoring the American College of Medical Genetics and Genomics and the Association for Molecular Pathology Guidelines for the Interpretation of Sequenced Variants in the FBN1 Gene for Marfan Syndrome: Proposal for a Disease- and Gene-Specific Guideline. Circ Genom Precis Med 11, e002039. 10.1161/CIRCGEN.117.002039.

10. Yoon, E., Lee, J.K., Park, T.K., Chang, S.A., Huh, J., Kim, J.W., Kim, D.K., and Jang, J.H. (2023). Experience of reassessing FBN1 variants of uncertain significance by gene-specific guidelines. J Med Genet. 10.1136/jmg-2023-109433.

11. Le Goff, C., Mahaut, C., Wang, L.W., Allali, S., Abhyankar, A., Jensen, S., Zylberberg, L., Collod-Beroud, G., Bonnet, D., Alanay, Y., et al. (2011). Mutations in the TGFbeta binding-protein-like domain 5 of FBN1 are responsible for acromicric and geleophysic dysplasias. Am J Hum Genet 89, 7–14. 10.1016/j.ajhg.2011.05.012.

12. Loeys, B., De Backer, J., Van Acker, P., Wettinck, K., Pals, G., Nuytinck, L., Coucke, P., and De Paepe, A. (2004). Comprehensive molecular screening of the FBN1 gene favors locus homogeneity of classical Marfan syndrome. Hum Mutat 24, 140–146. 10.1002/humu.20070.

13. Meester, J.A.N., Peeters, S., Van Den Heuvel, L., Vandeweyer, G., Fransen, E., Cappella, E., Dietz, H.C., Forbus, G., Gelb, B.D., Goldmuntz, E., et al. (2022). Molecular characterization and investigation of the role of genetic variation in phenotypic variability and response to treatment in a large pediatric Marfan syndrome cohort. Genet Med 24, 1045–1053. 10.1016/j.gim.2021.12.015.

14. Clarissa’s story: Marfan Syndrome. (2024). https://www.genomicsengland.co.uk/patients-participants/stories/clarissa.

15. Pagnamenta, A.T., Yu, J., Evans, J., Twiss, P., Genomics England Research, C., Musculoskeletal Ge, C.M., Offiah, A.C., Wafik, M., Mehta, S.G., Javaid, M.K., et al. (2023). Conclusion of diagnostic odysseys due to inversions disrupting GLI3 and FBN1. J Med Genet 60, 505–510. 10.1136/jmg-2022-108753.

16. Racine, C., Callier, P., Touraine, R., Vitobello, A., Hanna, N., Arnaud, P., Jondeau, G., Boileau, C., Thauvin-Robinet, C., Creveaux, I., et al. (2024). De Novo Balanced Translocations Disrupting the FBN1 Gene Diagnosed by Genome Sequencing: An Uncommon Cause of Marfan Syndrome Modifying Genetic Counseling. Am J Med Genet A, e63923. 10.1002/ajmg.a.63923.

17. Bonaglia, M.C., Salvo, E., Sironi, M., Bertuzzo, S., Errichiello, E., Mattina, T., and Zuffardi, O. (2023). Case Report: Decrypting an interchromosomal insertion associated with Marfan’s syndrome: how optical genome mapping emphasizes the morbid burden of copy-neutral variants. Front Genet 14, 1244983. 10.3389/fgene.2023.1244983.

18. Turnbull, C., Scott, R.H., Thomas, E., Jones, L., Murugaesu, N., Pretty, F.B., Halai, D., Baple, E., Craig, C., Hamblin, A., et al. (2018). The 100 000 Genomes Project: bringing whole genome sequencing to the NHS. BMJ 361, k1687. 10.1136/bmj.k1687.

19. de Sainte Agathe, J.M., Filser, M., Isidor, B., Besnard, T., Gueguen, P., Perrin, A., Van Goethem, C., Verebi, C., Masingue, M., Rendu, J., et al. (2023). SpliceAI-visual: a free online tool to improve SpliceAI splicing variant interpretation. Hum Genomics 17, 7. 10.1186/s40246-023-00451-1.

20. Jaganathan, K., Kyriazopoulou Panagiotopoulou, S., McRae, J.F., Darbandi, S.F., Knowles, D., Li, Y.I., Kosmicki, J.A., Arbelaez, J., Cui, W., Schwartz, G.B., et al. (2019). Predicting Splicing from Primary Sequence with Deep Learning. Cell 176, 535–548 e524. 10.1016/j.cell.2018.12.015.

21. Pagnamenta, A.T., Yu, J., Walker, S., Noble, A.J., Lord, J., Dutta, P., Hashim, M., Camps, C., Green, H., Devaiah, S., et al. (2024). The impact of inversions across 33,924 families with rare disease from a national genome sequencing project. Am J Hum Genet 111, 1140–1164. 10.1016/j.ajhg.2024.04.018.

22. Thomas, H.B., Wood, K.A., Buczek, W.A., Gordon, C.T., Pingault, V., Attie-Bitach, T., Hentges, K.E., Varghese, V.C., Amiel, J., Newman, W.G., and O’Keefe, R.T. (2020). EFTUD2 missense variants disrupt protein function and splicing in mandibulofacial dysostosis Guion-Almeida type. Hum Mutat 41, 1372–1382. 10.1002/humu.24027.

23. Bycroft, C., Freeman, C., Petkova, D., Band, G., Elliott, L.T., Sharp, K., Motyer, A., Vukcevic, D., Delaneau, O., O’Connell, J., et al. (2018). The UK Biobank resource with deep phenotyping and genomic data. Nature 562, 203–209. 10.1038/s41586-018-0579-z.

24. C Li, S., Carss, K.J., Halldorsson, B.V., Cortes, A., and Consortium, U.B.W.-G.S. (2023). Whole-genome sequencing of half-a-million UK Biobank participants. medRxiv, 2023.2012.2006.23299426. 10.1101/2023.12.06.23299426.

25. Gillis, E., Kempers, M., Salemink, S., Timmermans, J., Cheriex, E.C., Bekkers, S.C., Fransen, E., De Die-Smulders, C.E., Loeys, B.L., and Van Laer, L. (2014). An FBN1 deep intronic mutation in a familial case of Marfan syndrome: an explanation for genetically unsolved cases? Hum Mutat 35, 571–574. 10.1002/humu.22540.

26. Ogawa, N., Imai, Y., Takahashi, Y., Nawata, K., Hara, K., Nishimura, H., Kato, M., Takeda, N., Kohro, T., Morita, H., et al. (2011). Evaluating Japanese patients with the Marfan syndrome using high-throughput microarray-based mutational analysis of fibrillin-1 gene. Am J Cardiol 108, 1801–1807. 10.1016/j.amjcard.2011.07.053.

27. Calvo, S.E., Pagliarini, D.J., and Mootha, V.K. (2009). Upstream open reading frames cause widespread reduction of protein expression and are polymorphic among humans. Proc Natl Acad Sci U S A 106, 7507–7512. 10.1073/pnas.0810916106.

28. Golledge, J. (2019). Abdominal aortic aneurysm: update on pathogenesis and medical treatments. Nat Rev Cardiol 16, 225–242. 10.1038/s41569-018-0114-9.

29. Guo, D.C., Gupta, P., Tran-Fadulu, V., Guidry, T.V., Leduc, M.S., Schaefer, F.V., and Milewicz, D.M. (2008). An FBN1 pseudoexon mutation in a patient with Marfan syndrome: confirmation of cryptic mutations leading to disease. J Hum Genet 53, 1007–1011. 10.1007/s10038-008-0334-7.

30. Kim, J.A., Jang, M.A., Jang, S.Y., Kim, D.K., Kim, Y.G., Kim, J.W., Park, T.K., and Jang, J.H. (2024). Overcoming challenges associated with identifying FBN1 deep intronic variants through whole-genome sequencing. J Clin Lab Anal 38, e25009. 10.1002/jcla.25009.

31. Smith, C., and Kitzman, J.O. (2023). Benchmarking splice variant prediction algorithms using massively parallel splicing assays. Genome Biol 24, 294. 10.1186/s13059-023-03144-z.

32. Hiraide, T., Shimizu, K., Miyamoto, S., Aoto, K., Nakashima, M., Yamaguchi, T., Kosho, T., Ogata, T., and Saitsu, H. (2022). Genome sequencing and RNA sequencing of urinary cells reveal an intronic FBN1 variant causing aberrant splicing. J Hum Genet 67, 387–392. 10.1038/s10038-022-01016-1.

33. Rodriguez-Munoz, A., Liquori, A., Garcia-Bohorquez, B., Jaijo, T., Aller, E., Millan, J.M., and Garcia-Garcia, G. (2022). Functional assays of non-canonical splice-site variants in inherited retinal dystrophies genes. Sci Rep 12, 68. 10.1038/s41598-021-03925-1.

34. Miller, D.T., Lee, K., Abul-Husn, N.S., Amendola, L.M., Brothers, K., Chung, W.K., Gollob, M.H., Gordon, A.S., Harrison, S.M., Hershberger, R.E., et al. (2023). ACMG SF v3.2 list for reporting of secondary findings in clinical exome and genome sequencing: A policy statement of the American College of Medical Genetics and Genomics (ACMG). Genet Med 25, 100866. 10.1016/j.gim.2023.100866.

35. Robinson, D.O., Lin, F., Lyon, M., Raponi, M., Cross, E., White, H.E., Cox, H., Clayton-Smith, J., and Baralle, D. (2012). Systematic screening of FBN1 gene unclassified missense variants for splice abnormalities. Clin Genet 82, 223–231. 10.1111/j.1399-0004.2011.01781.x.

36. De Angeli, P., Flores-Tufino, A., Stingl, K., Kuhlewein, L., Roschi, E., Wissinger, B., and Kohl, S. (2024). Splicing defects and CRISPR-Cas9 correction in isogenic homozygous photoreceptor precursors harboring clustered deep-intronic ABCA4 variants. Mol Ther Nucleic Acids 35, 102113. 10.1016/j.omtn.2023.102113.

37. Luo, X., Wang, R., Sun, Y., Qiu, W., Lu, D., Wang, Y., Gong, Z., Zhang, H., Han, L., Liang, L., et al. (2023). Deep Intronic PAH Variants Explain Missing Heritability in Hyperphenylalaninemia. J Mol Diagn 25, 284–294. 10.1016/j.jmoldx.2023.02.001.

38. Oh, R.Y., AlMail, A., Cheerie, D., Guirguis, G., Hou, H., Yuki, K.E., Haque, B., Thiruvahindrapuram, B., Marshall, C.R., Mendoza-Londono, R., et al. (2024). A systematic assessment of the impact of rare canonical splice site variants on splicing using functional and in silico methods. HGG Adv 5, 100299. 10.1016/j.xhgg.2024.100299.

39. Lerner-Ellis, J.P., Aldubayan, S.H., Hernandez, A.L., Kelly, M.A., Stuenkel, A.J., Walsh, J., and Joshi, V.A. (2014). The spectrum of FBN1, TGFbetaR1, TGFbetaR2 and ACTA2 variants in 594 individuals with suspected Marfan Syndrome, Loeys-Dietz Syndrome or Thoracic Aortic Aneurysms and Dissections (TAAD). Mol Genet Metab 112, 171–176. 10.1016/j.ymgme.2014.03.011.

40. Overwater, E., Floor, K., van Beek, D., de Boer, K., van Dijk, T., Hilhorst-Hofstee, Y., Hoogeboom, A.J.M., van Kaam, K.J., van de Kamp, J.M., Kempers, M., et al. (2017). NGS panel analysis in 24 ectopia lentis patients; a clinically relevant test with a high diagnostic yield. Eur J Med Genet 60, 465–473. 10.1016/j.ejmg.2017.06.005.

