## Supplemental for "Utility of genome sequencing and group-enrichment to support splice variant interpretation in Marfan syndrome"

***Supplementary Methods***

***Further details of bioinformatics analysis***

The 100,000 Genomes Project (100kGP) is a nationwide study that aimed to uncover the genetic basis of disease for individuals in whom a diagnosis had not been obtained by standard-of-care testing.^1^ Data from the 100kGP is held in the National Genomic Research Library, Genomics England, <https://doi.org/10.6084/m9.figshare.4530893.v7>). Version 10 of the 100kGP release was used for a large-scale aggregation that included germline data for 78,195 participants from both the rare disease and cancer programmes of the 100kGP. This dataset (the “AggV2” file) includes >722 million SNVs and small indels (≤50 bp) and is split into 1,371 genome chunks of approximately equal size, with the one containing *FBN1* corresponding to a 2.81Mb segment on 15q21.1 (chr15:46,244,291-49,055,668, GRCh38). Genome sequencing was using a 150bp paired-end read format in a single lane of an HiSeqX instrument (Illumina). Data was uniformly processed on the Illumina North Star Version 4 Whole Genome Sequencing Workflow (NSV4, v2.6.53.23); which comprises the iSAAC Aligner (v03.16.02.19) and Starling Small Variant Caller (v2.4.7). Reads were aligned to the NCBI GRCh38 assembly with decoys. Singleton variants were extracted from this AggV2 file using BCFtools v.1.16 for the entire *FBN1* gene region using the GRCh38 coordinates: chr15:48,408,313-48,645,709. Variant annotation was performed using VEP v99.1. Odds Ratios (ORs) and 2-tailed significance levels based on Fisher’s exact test were calculated using the 2024.04.2+764 version of RStudio.

***Recruitment criteria for Familial Thoracic Aortic Aneurysm Disease***

For Families to be recruited to the 100kGP under a diagnosis of Familial Thoracic Aortic Aneurysm Disease (FTAAD), patients had to be suspected to have one of the following conditions: Familial Thoracic Aortic Aneurysm and dissection, Thoracic aortopathy at < 50 years with no other established risk factors, clinically diagnosed Marfan syndrome with no *FBN1* mutation, Loeys-Dietz syndrome and Loeys-Dietz syndrome like conditions, mutation negative Congenital Contractural Arachnodactyly (Beals syndrome).

Families where there were other potential (i.e. non-genetic) risk factors for sporadic thoracic aortopathies were excluded. Another criteria for exclusion was for cases with family history but where there were no affected probands to test. Prior genetic testing should have been undertaken for any individual gene for which diagnostic yield is >10% and this includes Loeys-Dietz syndrome (*TGFBR1* and *TGFBR2*), Marfan Syndrome (*FBN1*), Congenital Contractural Arachnodactyly (*FBN2*) and Isolated familial thoracic aortic aneurysms and dissection (*ACTA2*). The above requirements were in place at the start of the 100kGP but were kept under continual review during the main programme.

***RNAseq***

Short read RNAseq had already been performed for 10 of the 23 families with a candidate splice variant in *FBN1*, of which 8 were from the 100kGP pilot transcriptomics project, one from the NHS transformation project and the last performed at the Manchester Centre for Genomic Medicine. RNAseq data generated as part of Genomics England’s Transcriptomics Pilot study comprised RNA-sequencing of 5546 rare disease probands from the 100kGP who initially did not receive a genetic diagnosis through the Genomics England primary pipeline. Priority for inclusion in this cohort was based on a number of factors that included: i) individuals with variants highlighted through SpliceAI, ii) autosomal recessive disorders with only a single pathogenic variant identified, iii) VUS with either high Exomiser score or selected by the local Genomic Medicine Centre and iv) conditions that are considered highly likely to have a monogenic cause. Total RNA was from peripheral whole blood prepared using QIAGEN PAXgene Blood RNA Kit. Integrity of the isolated total RNA was assessed on an Agilent Tapestation 4200 system. Libraries were constructed following the Illumina Stranded Total RNA Prep, Ligation with Ribo-Zero Plus kit protocol. Sequencing was done with 95 samples per plate (one empty well) simultaneously on the NovaSeq 6000 using 2x100bp paired-end reads. The Illumina DRAGEN RNA Pipeline (v3.8.4) was used for read alignment. Reads were mapped to GRCh38 with annotation-assisted alignment, duplicate marking, gene expression quantification, and gene fusion detection enabled.

***Targeted RNA analysis***

For seven of the twenty variants, RNA testing was done by the Wessex Genomics Laboratory Service, using a well-established protocol optimised for low abundance transcripts. Blood samples were collected into PAXgene RNA collection tubes and RNA was extracted using the QIAcube Connect extraction machine (Qiagen, UK) and the RNeasy kit (Qiagen, UK) followed by cDNA preparation using 1.5µl of 10mM dNTP mix (Promega, UK), 2µl of 0.1M Dithiothreitol (Invitrogen, UK), 1µl of 40 U/µl Moloney Murine Leukemia Virus Reverse Transcriptase (Invitrogen, UK), 1µl of 40 U/µl RNAaseOut ribonuclease inhibitor (Invitrogen, UK), 4µl of Reverse Transcriptase buffer (Invitrogen, UK), 1µl of 10ng/µl random hexamer primers (Thermo Fisher, UK) and 10µl of RNA. Samples were incubated for one hour at 37°C followed by 10mins at 65°C.

RT-PCR reactions were carried out in a 20µl volume containing 1.5µl of cDNA, 10nM of each primer (Thermo Fisher, UK), 2µl of 10x Platinum Taq buffer (Invitrogen, UK), 0.2mM of each dNTP, 1.5mM MgCl_2_ and 0.5 units of Platinum Taq polymerase (Invitrogen, UK). Cycling parameters were 94°C for 12 minutes followed by 35 cycles of 94°C for 30 seconds, 60°C for 30 seconds and 72°C for 30 seconds. Primer sequences are listed in Table S3. PCR products were checked by gel electrophoresis and then bi-directionally sequenced using the standard protocol of the BigDye Terminator v1.1 Cycle Sequencing Kit (Applied Biosystems, USA) and separated on an ABI 3130xl Genetic Analyzer (Applied Biosystems, USA). Subsequent data were analysed using the Mutation Surveyor (version 3.1) software (SoftGenetics, USA).

For Families 3/17, similar methodologies to those described above were employed, except that for Family 3 PCRs were with MyFi Master mix (Bioline) and performed in 10µl volumes. In contrast, for Family 17 RT-PCRs used RNA obtained from fibroblasts ±CHX treatment, to monitor the effects of NMD.

**Comparison of costs and turnaround time for targeted vs minigene testing**

For RT-PCRs performed by the Wessex Genomics Laboratory Service, the official turnaround time is 6 weeks. RNA extraction takes about half a day and the PCR/sequencing takes about two days. Generally, not much optimisation is required, but designing bespoke primers for deep intronic variants can be tricky as was seen for Family 7. The RNA extraction is typically the bottleneck and samples average two weeks for that step plus one week for post-extraction work, so in practise, the average turnaround time is three weeks per sample. Approximate costs and hands-on time are shown below.

|  | **Time/sample (hours)** | **Cost (£/sample)** | **Notes** |
| --- | --- | --- | --- |
| PAXgene tubes | - | 25 | - |
| RNA extraction | 1 | 5 | Can do six at once |
| cDNA synthesis | 1 | 10 | Can do six at once in 1 hour |
| Primers | 1 | 20 | Can do six at once in 1 hour |
| Agarose gel | 1 | 5 | Can do all on run in 1 hour |
| Sequencing | 1 | 15 | Can do all on run in 1 hour |
| Analysis/Reporting | 1 | - | - |
| Total | 6 | 80 | - |

The whole minigene assay is multi-step experiment and necessitates a slightly higher degree of expertise in molecular biology. However, all steps are relatively straightforward and cheap but often 1 or more steps need repeating. Turnaround time needs to factor in the time taken for cells to grow and for Sanger sequencing results to be returned and so the time from design to completion can be anything between 2-6 weeks. Multiplexing is possible such that up to 4 variants can be assayed easily at the same time (against the same WT), but the size of minigene construct and number of exons included in the inserted sequence typically add a little to the cost and likely timescales. Overall, £150 is a good approximation on costs per assay (for both the WT and variant construct) but these can vary widely. A breakdown of the approximate costs and hands-on time are shown below.

|  | **Time/sample (hours)** | **Cost (£/sample)** |
| --- | --- | --- |
| Primer design and PCR amplification of minigene | 2 | 15 |
| Vector cloning | 1 | 25 |
| Bacterial transformation | 1 | 5 |
| Vector isolation (and Sanger sequencing) | 2 | 40 |
| HEK cell transfection | 2 | 5 |
| RNA extraction | 3 | 20 |
| cDNA synthesis | 1 | 5 |
| RT-PCR (+ Sanger sequencing) | 2 | 40 |
| Total | 14 | 145 |

***Supplementary Note: haplotype studies and gnomAD data for c.1589-1217G>T***

Haplotype studies for Families 12-15 were limited by the fact that only 2/4 families (Families 12 and 13) were analysed on GRCh38 whilst the other two were on GRCh37. Using the families on GRCh38 we performed several analyses that included: i) conflicting homozygosity analysis using common high quality SNVs, ii) searches for shared ultra-rare variants (<0.1%) using bcftools and the AggV2 dataset and iii) comparison of the 100kGP data to a set of 16 rare SNVs found on the disease haplotype in the original TAA758 family (pers. comm. Dong-Chuan Guo).^2^

Although c.1589-1217G>T is reported in gnomAD at an allele frequency of 2/147,744, for both heterozygous individuals the alignments looks suspicious - the variant is only seen on -ve strand reads and it is flagged as failing the allele-specific VQSR filter. In contrast, read alignments for the individuals from the 100kGP indicated a higher quality variant call (Figure S10). Review of the GRCh38 reference genome sequence shows that this variant (15:48511386C>A) is immediately adjacent to a run of nine As. Although this long homopolymer may explain the presence of artifactual calls in gnomAD, it could also precipitate genuine mutational reccurence.

***Supplementary Figures***

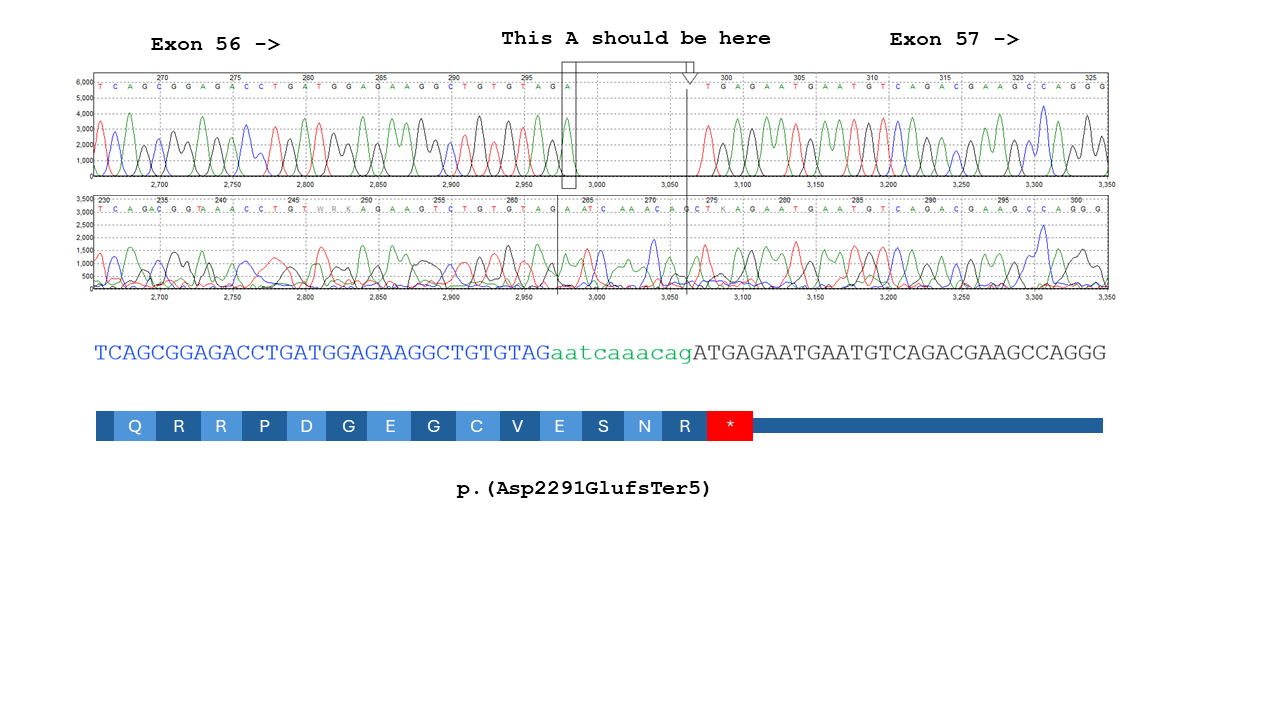

**Figure S1:** Sanger sequencing of RT-PCR product for the proband from Family 1 showing a 10bp extension at the 5’ end of exon 57, r.6871_6872insaaucaaacag (green) and predicting p.(Asp2291GlufsTer5). These results were consistent with the SpliceAI predictions for NM_000138.5:c.6872-11A>G which were for a near-perfect switch of acceptor sites (AG=1.00, -1; AL=0.99, -11).

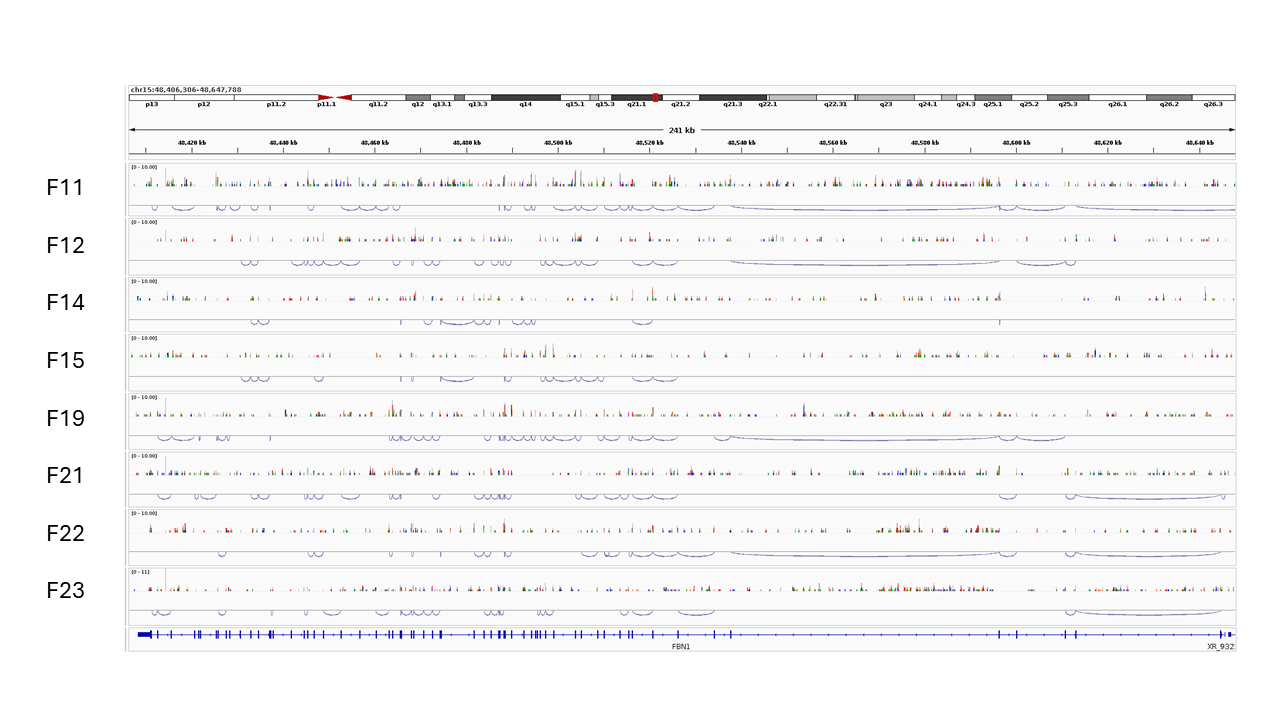

**Figure S2:** Full gene view showing RNAseq data across 8 of the 23 families in the present study for whom this data was available as part of the Genomics England Pilot RNAseq project. The sparse coverage is as expected, given the low expression of *FBN1* in blood.

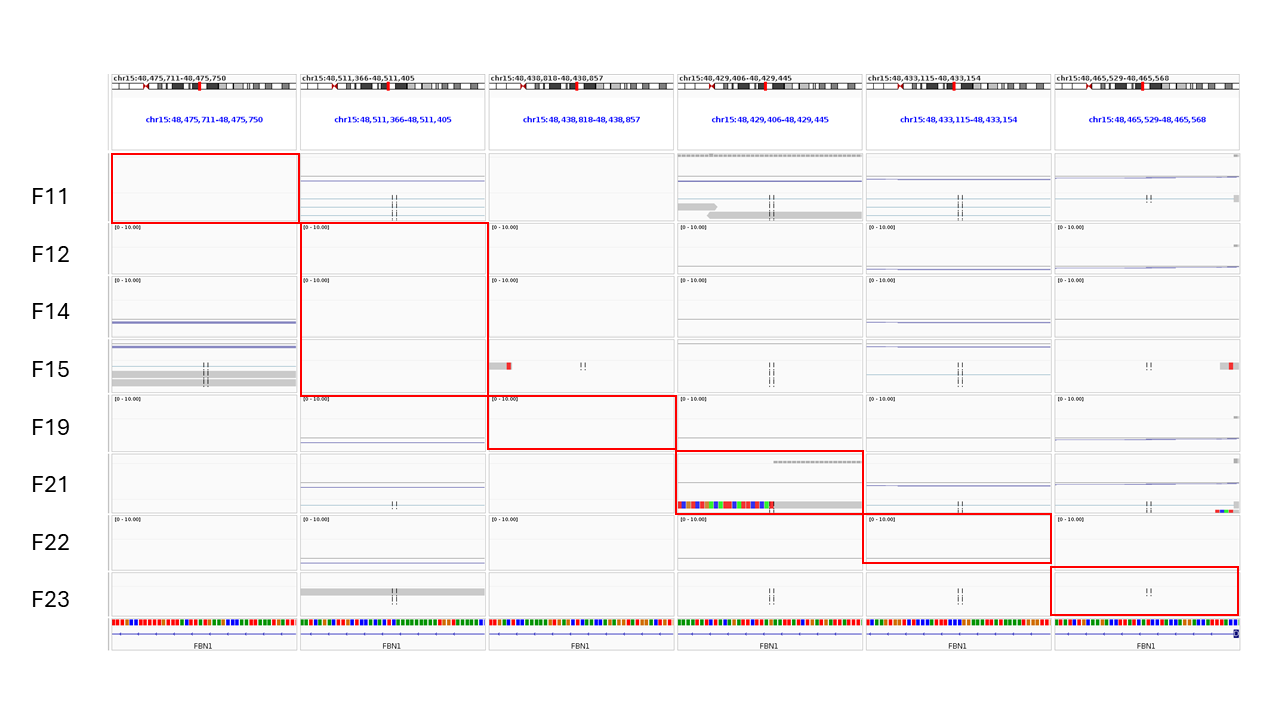

**Figure S3:** Multi-region view where the six 41bp IGV windows show intronic regions of interest. Red rectangles highlight the relevant locus for each family, with F12, F14 and F15 all harbouring the same recurrent c.1589-1217G>T variant. Across these eight datasets, only a single read was observed at an informative position and this spanned the c.6872-955C>G variant in F21. The soft clipped sequence corresponds to the junction between the predicted 96bp pseudoexon and the start of exon 57.

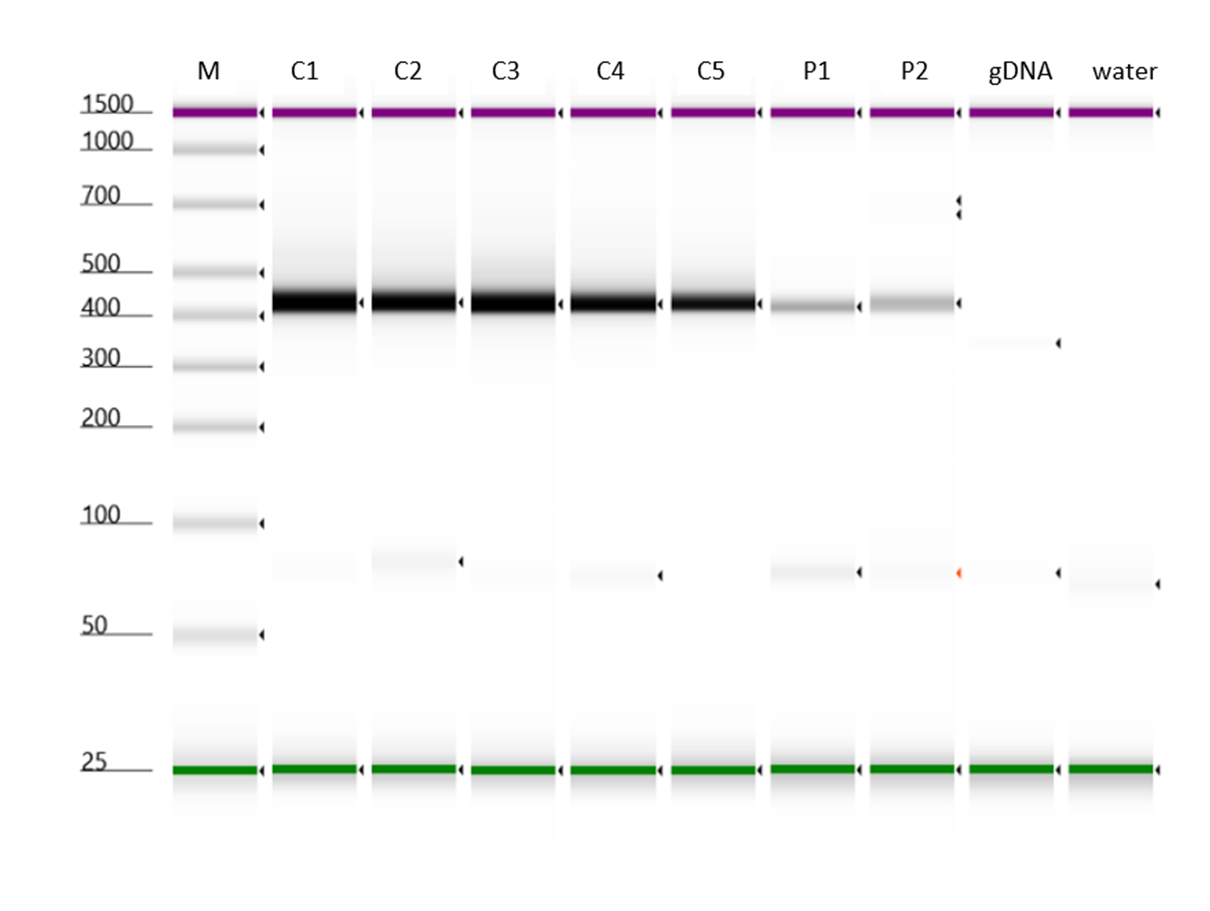

**Figure S4:** RT-PCR products for Family 3 suggest a possible nonsense mediated decay effect associated with the c.6038-14T>G variant. PCRs were performed with N13 tagged primers using cDNA generated from five unaffected controls (C1-5), with the patient being tested in duplicate (P1, P2). A gDNA control and a water blank were included as negative controls. 1µL of each of the PCR was analysed on the Tapestation 4200 using a D1000 screentape (Agilent). No difference in size was observed in the amplicon sizes and Sanger sequencing further confirmed no qualitative differences.

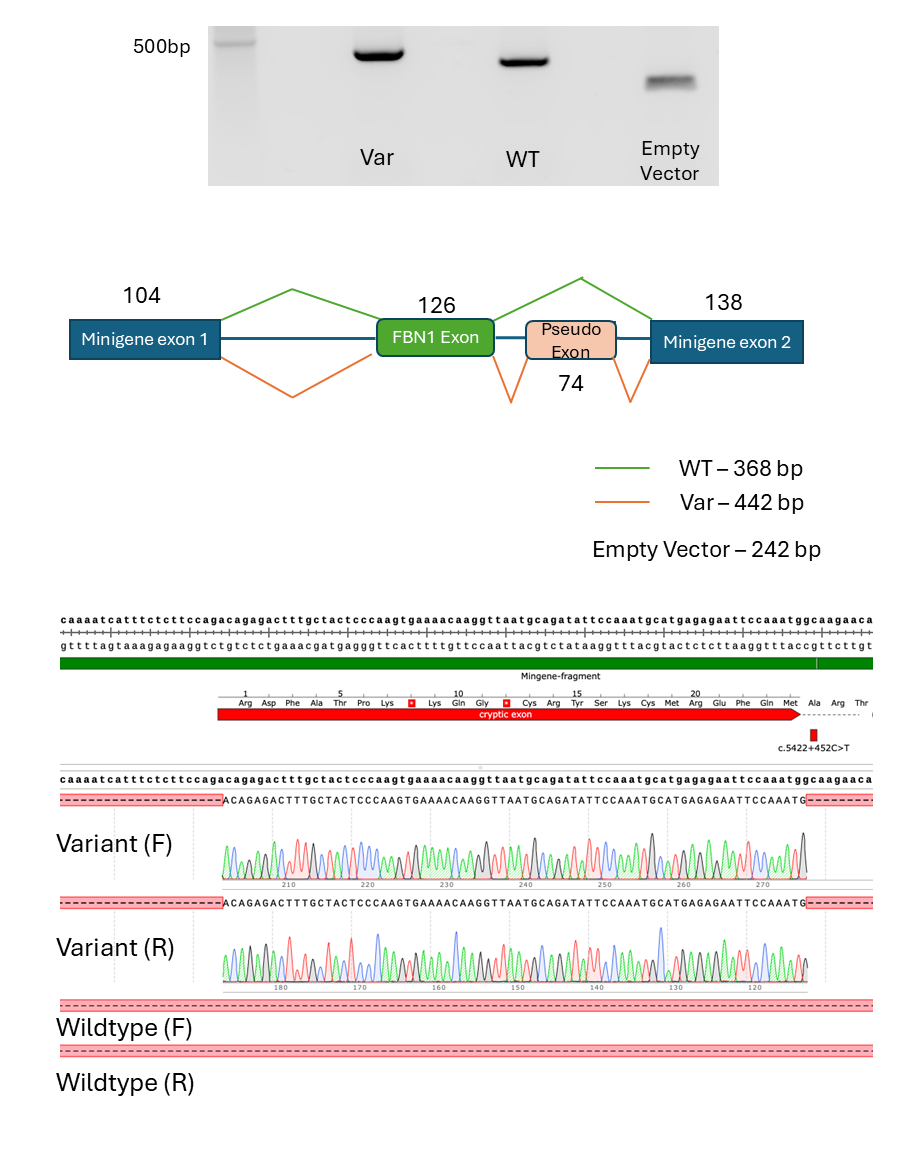

**Figure S5:** Minigene test and Sanger sequencing Family 16 supports the *in silico* prediction for c.5422+452C>T. This heterozygous variant was identified in both the affected father and daughter. Upper image show agarose gel showing increased size of band for the variant, consistent with inclusion of a 74bp pseudoexon and the SpliceAI predictions in for a donor gain (DG=0.94, +2) and an acceptor gain (AG=0.83, +75). Sanger sequencing confirmed the results and electropherograms are shown for both directions. The inserted sequence r.5422_5423ins74 was not observed for the wildtype construct and is predicted to result in 7 novel amino acids followed by a premature termination codon p.(Ile1809ArgfsTer8).

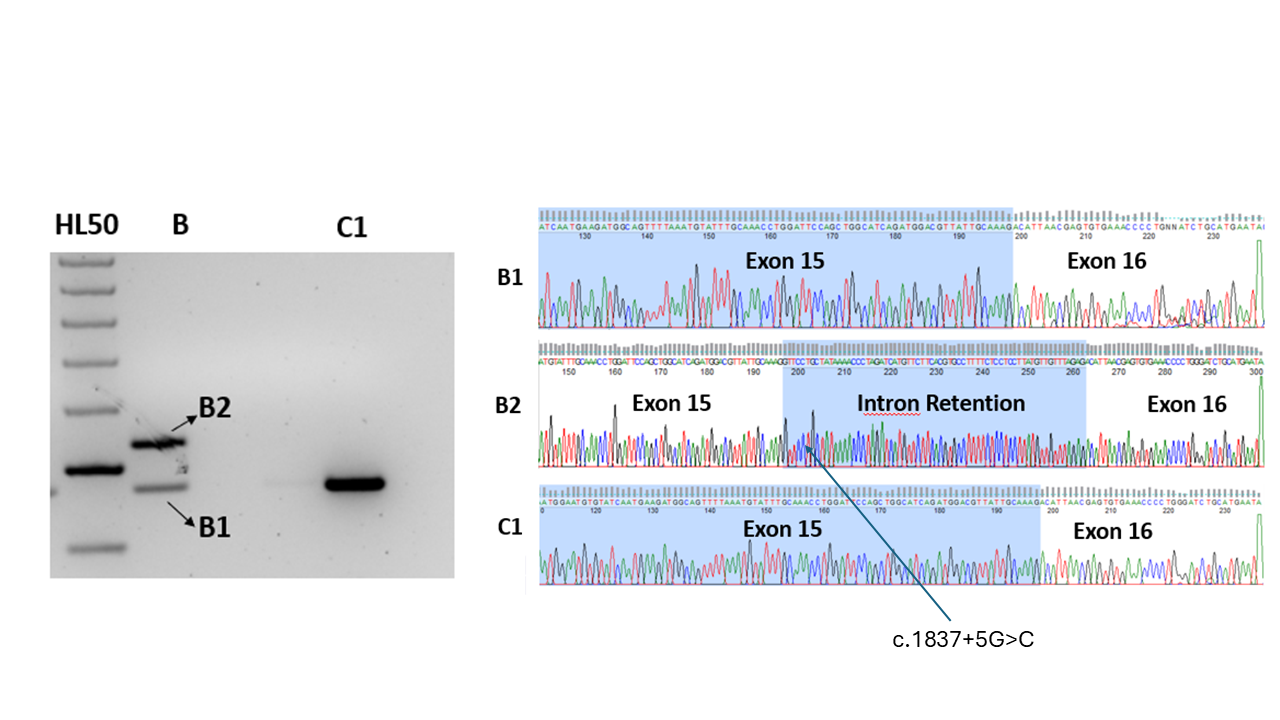

**Figure S6:** Gel electrophoresis and Sanger sequencing of RT-PCR products obtained for the son in Family 17. Bands 1 (B1) and 2 (B2) were excised from the gel and sequenced alongside a control sample (C1). The c.1837+5G>C variant which only has a moderate score for a donor loss (DL=0.38). RT-PCR from blood confirmed an in frame 66bp extension of exon 15. The position of c.1837+5G>C is labelled.

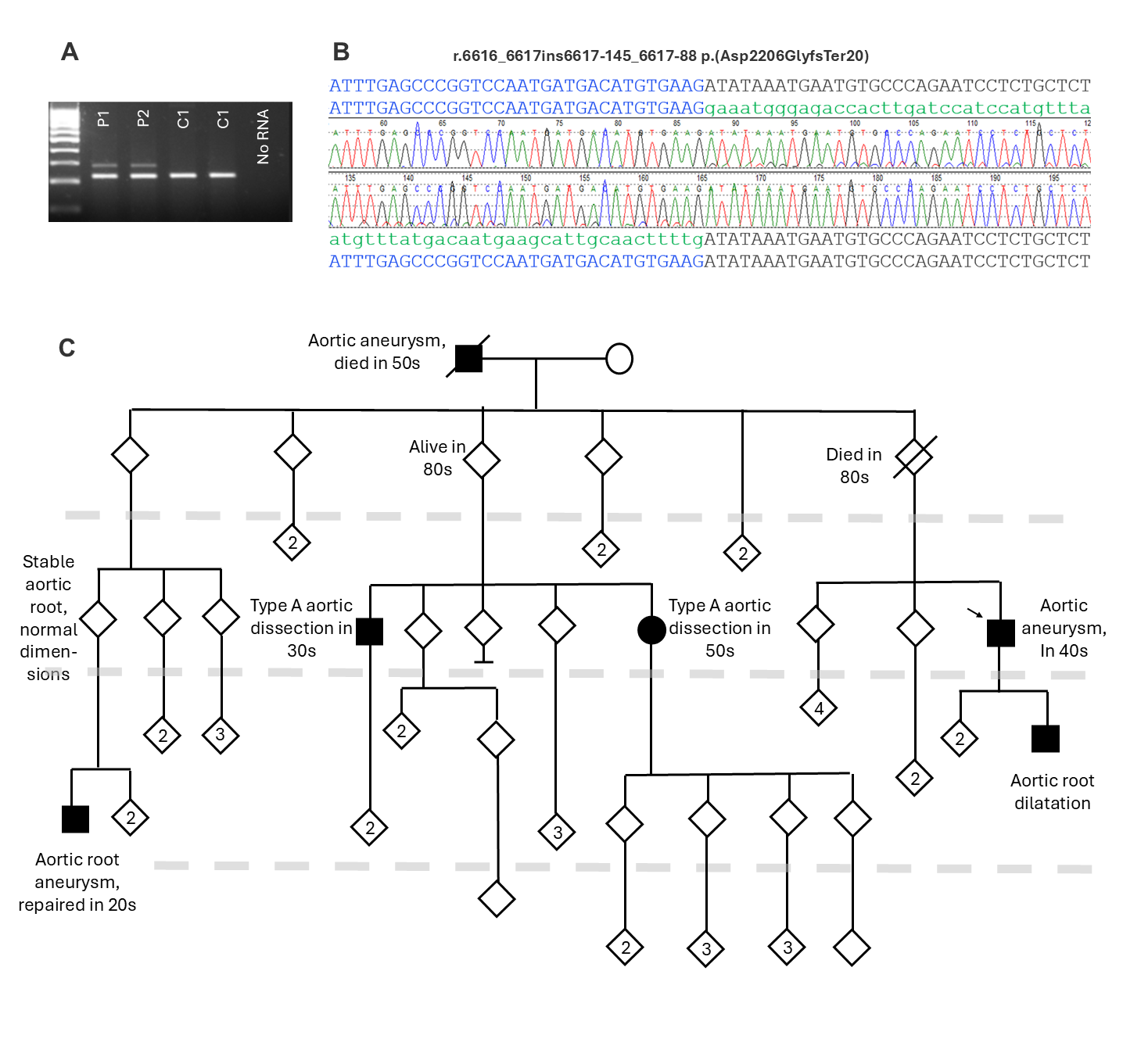

**Figure S7:** A) Gel electrophoresis and B) Sanger sequence data for Family 22 confirming effect of the c.6617-147G>A variant. The variant has a moderate prediction for an acceptor gain (AG=0.44, -2) but with the larger window, there is also a stronger prediction for a donor gain (DG=0.57, -59). Consistent with these *in silico* results, RT-PCR and Sanger sequencing from blood RNA confirmed the presence of a pseudoexon, r.6616_6617ins6617-145_6617-88, p.(Asp2206GlyfsTer20). The upper band on the gel is a lot weaker and so the Sanger trace corresponding to the inserted sequence (green), although clearly visible, is at a much lower intensity. C) Pedigree for Family 22, with multiple affected branches and a large number of at-risk individuals who could benefit from cascade testing. The presence of at least 2 obligate carriers unaffected into their 80s highlights the incomplete penetrance seen for this condition. Generations are separated by a dotted grey horizontal line. Diamond symbols are used to hide gender for unaffected/undiagnosed individuals.

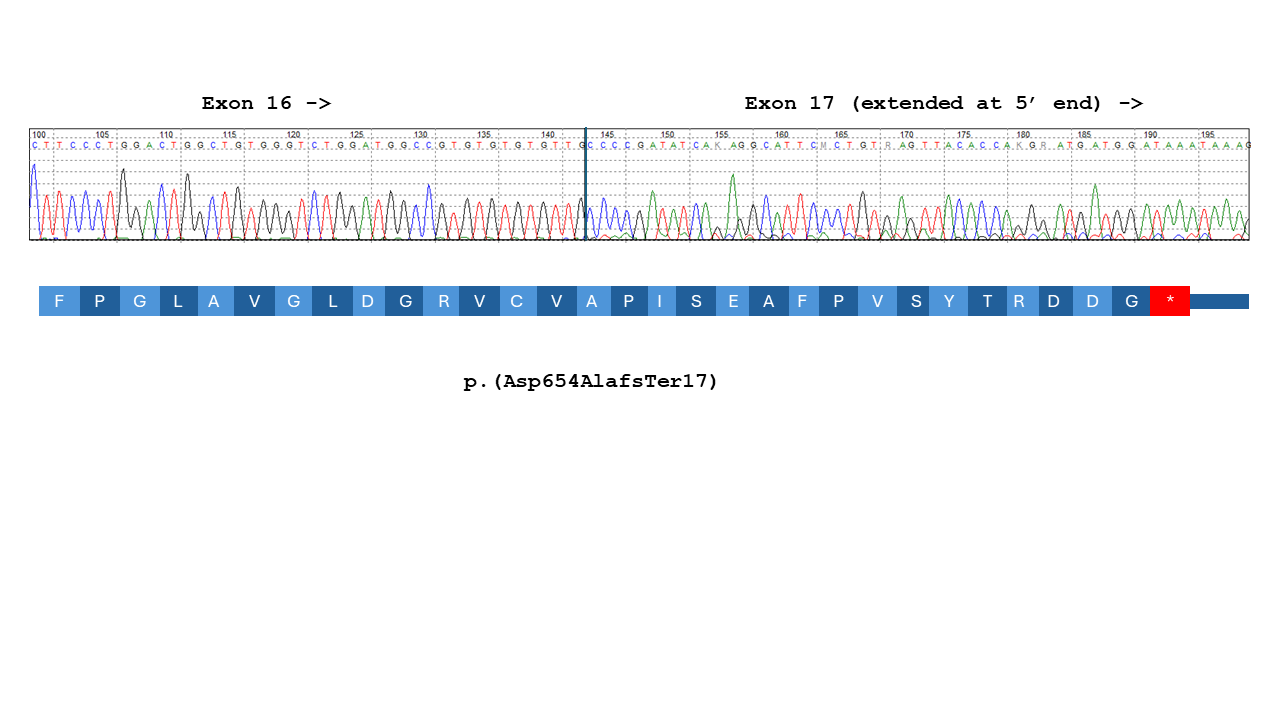
**Figure S8:** Sanger sequencing of main RT-PCR product for the proband from Family 7 showing a 133bp extension of exon 17 and predicting p.(Asp654AlafsTer17). Standard RT-PCR primers had previously failed to detect this large insertion and it was only a combination of group enrichment and *in silico* predictions with the larger analysis window that prompted reanalysis with customised primers.

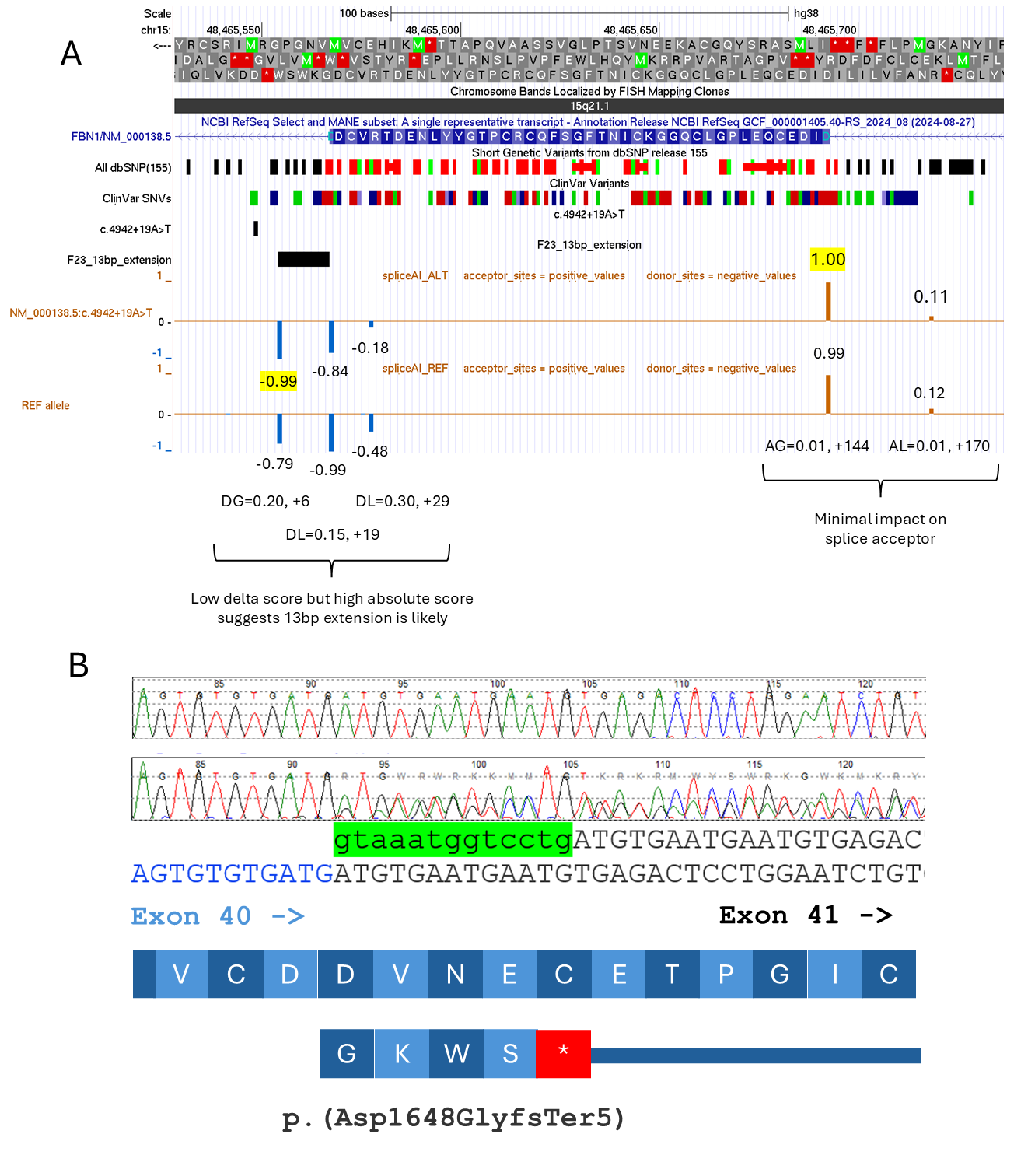

**Figure S9:** Variant in Family 23 highlights importance of assessing both relative and absolute SpliceAI scores. A) UCSC genome browser graphic showing the SpliceAI scores for c.4942+19A>T in comparison to the reference allele. The highest absolute scores are highlighted in yellow and strongly suggest extension of exon 40 by 13bp. B) PCR-Sanger sequencing of cDNA indicates a 13bp insertion corresponding to the sequence from the extended exon (highlighted in green) that is predicted to lead to a frameshift and termination after the insertion of 4 novel amino acids.

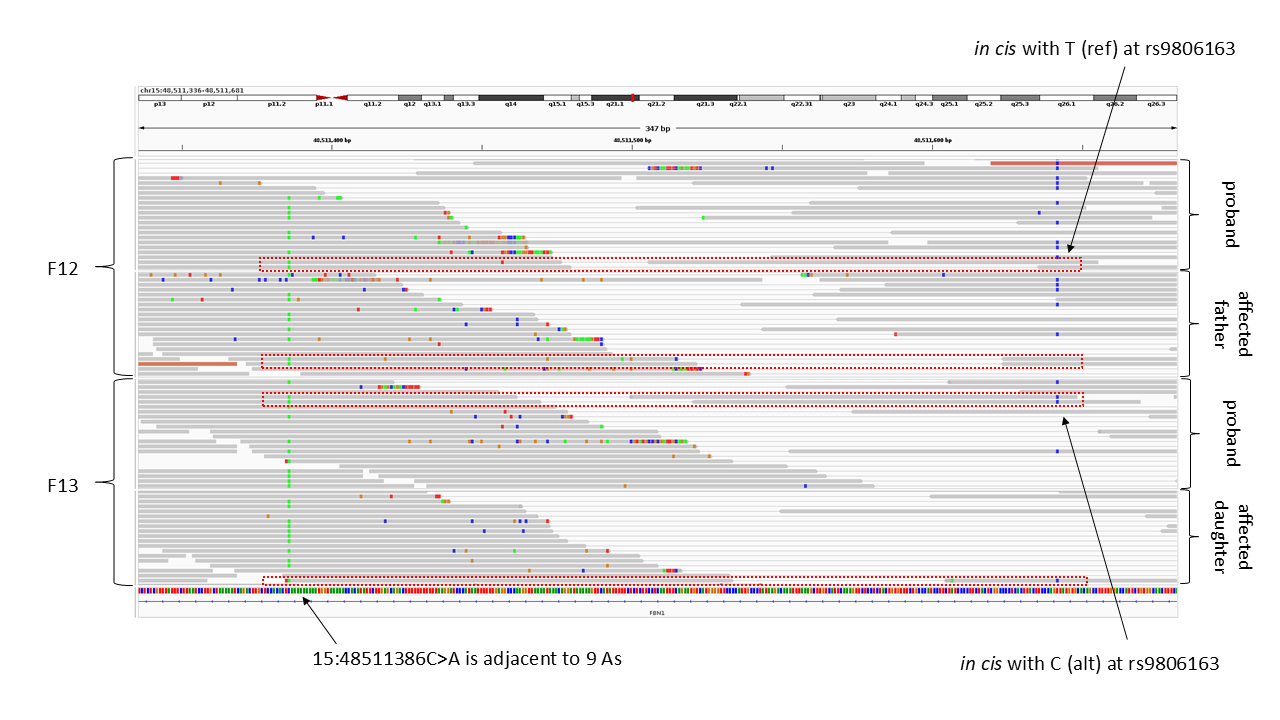
**Figure S10:** Paired-read level data for 4 individuals from Families 12 and 13 indicate that c.1589-1217G>T resides on two distinct haplotypes and implies independent mutational recurrence. Informative read-pairs that capture both the pathogenic variant and the common variant 256bp away are highlighted by red boxes. Phasing was not possible by transmission as all sequenced family members were heterozygous. Data for Families 14 and 15 was analysed on GRCh37 and so is not shown, although for both these families, c.1589-1217G>T is *in cis* with the T allele at rs9806163. The pathogenic variant (15:48511386C>A , GRCh38) lies next to a run of 9As and this likely explains the mutational recurrence.

***Supplementary Tables***

**Table S1:** Examples of previously published intronic variants in *FBN1* outside ±8bp splice region. GS, genome sequencing; AS_VQSR, failed allele-specific variant quality score recalibration filter; CHX, cyclohexamide; MFS, Marfan syndrome. †Incorrectly reported as IVS63+373, as “acceptor gain” and that first codon of pseudoexon is a stopgain. ‡, 35 genes, see [www.ncbi.nlm.nih.gov/gtr/tests/530671](http://www.ncbi.nlm.nih.gov/gtr/tests/530671).

| **cDNA annotation (NM_000138.5)** | **GRCh38 coordinates** | **SpliceAI prediction (delta position)** | **Effect on RNA/protein (NP_000129.3)** | **Variant detection method(s)** | **RNA confirmation method** | **gnomAD v4.1.0** | **Other comments** | **ClinVar** | **Publication** |
| --- | --- | --- | --- | --- | --- | --- | --- | --- | --- |
| c.8051+375G>T | 15:48415161C>A | DG=0.31 (2)† | 93bp pseudoexon (48415163-48415255)  p.(G2684_H2685insDSS LCKYTSLPTHDRTGLTLHTTDPLFSWGR) | Sanger sequencing of intron 63 | Sequencing of cDNA from fibroblasts and pyrosequencing | Absent | First report of *FBN1* pseudoexon  leading to MFS. | VCV000406327.4 | Guo *et al* 2008^3^ |
| c.6872-961A>G | 15:48429432T>C | DG=0.97 (1) | 90bp pseudoexon (48429433-48429522) containing stop codon p.(Asp2291ValfsTer9) | Linkage and Sanger sequencing of intron 56 | Sequencing of cDNA from fibroblasts with/without puromycin | Absent | Large 5 generation family | VCV000643480.3 | Gillis *et al* 2014^4^ |
| c.6872-24T>A | 15:48428495A>T | AG=1.00 (-2) and AL=0.99 (-24) | Extends exon by 22bp r.6871_6872ins6872-22_6872-1 and p.(Asp2291GlyfsTer9) | PCR-Sanger | RT-PCR using RNA extracted from blood lymphocyte fraction using RNeasy Mini Kit | Absent | Co-segregation in affected father. Evidence of NMD | NA | Fusco *et al* 2019^5^ |
| c.7571-12T>A | 15:48421698A>T | AG=0.99 (-2) and AL=0.96 (-12) | Extends exon by 10bp (r.7570_7571ins7571-10_7571-1) and p.(Asp2524ArgfsTer5) | NGS using HaloPlex gene panel and MiSeq |  | Absent | No other family members available for testing | NA |  |
| c.2678-15C>A | 15:48494269G>T | AG=0.99 (-2) and AL=0.87 (-15) | 13bp extension of exon 23 p.(Asp893AlafsTer10) | Targeted NGS of aortic disease panel‡ | Minigene in HeLa and COS-7 cells | Absent |  | VCV000446494.4 | Torrado *et al* 2018^6^ |
| c.1589-1217G>T | 15:48511386C>A | AG=0.20 (-3) | 202bp pseudoexon (48511182-48511383) containing stop codon p.(Asp530ValfsTer8) | Linkage and GS | Sequencing of cDNA from fibroblasts with/without CHX and qRT-PCR | 2/147744 (AS_VQSR filter) | Large 5 generation family | VCV002664316.1 | Guo *et al* 2023^2^ |
| c.5789-15G>A | 15:48445519C>T | AL=0.24 (-15) | Retention of intron 47 p.(Asp1930GlyfsTer20) | GS | RNAseq and RT-PCR in urinary cells. Allelic imbalance using p.N625N | Absent | Argues for urine being used for RNA studies. | NA | Hiraida *et al* 2022^7^ |
| c.6163+1484A>T | 15:48440237T>A | DG=0.87 (2), AG=0.69 (192) | Pseudoexons of 99bp and 187bp in intron 50, the longer of which has PTC p.(Asp2055AlafsTer7) | GS | RT-PCR using lymphocyte RNA | Absent | Prediction of cryptic splice acceptor by SpliceAI was 4bp out | NA | Kim *et al* 2024^8^ |
| c.5788+36C>A | 15:48446670G>T | DG=0.97 (3) | Retention of 33bp between exons 47/48. Predicted to add 11 amino acids, p.(D930delinsGACAKLCISKGN) in cbEGF domain | GS | RT-PCR using fibroblast RNA | Absent | NA | VCV000549305.1 |  |

**Table S2:** Summary of top 15 most common diagnostic categories for individuals contained within the aggregate dataset (“AggV2”) in the 100k Genomes Project. The total number of individuals in this cohort is 78,195. Ignoring the N/A category which mostly comprises unaffected family members, FTAAD is the 11^th^ most common diagnostic category in this cohort, but overall this represents less than 1% of individuals in AggV2.

| Rank | Normalised specific disease | Number in AggV2 | Percentage in diagnostic category |
| --- | --- | --- | --- |
| 1 | N/A | 28,197 | 36.06% |
| 2 | Cancer germline | 15,155 | 19.38% |
| 3 | Intellectual disability | 6275 | 8.02% |
| 4 | Cystic kidney disease | 1488 | 1.90% |
| 5 | Ultra-rare undescribed monogenic disorders | 1455 | 1.86% |
| 6 | Epilepsy plus other features | 1409 | 1.80% |
| 7 | Rod-cone dystrophy | 1107 | 1.42% |
| 8 | Hereditary ataxia | 1017 | 1.30% |
| 9 | Congenital Anomaly of the Kidneys and Urinary Tract | 907 | 1.16% |
| 10 | Hypertrophic cardiomyopathy | 807 | 1.03% |
| 11 | Familial breast and or ovarian cancer | 784 | 1.00% |
| *12* | ***Familial Thoracic Aortic Aneurysm Disease*** | **703** | **0.90%** |
| 13 | Charcot-Marie-Tooth disease | 648 | 0.83% |
| 14 | Congenital hearing impairment | 629 | 0.80% |
| 15 | Primary immunodeficiency | 614 | 0.79% |

**Table S3:** Primers used for RNA validation experiments, where available. *N13 tagged sequence in lower case. †Intronic primer for repeat RT-PCRs, based on SpliceAI prediction. ‡designed across variant breakpoint.

| **Family** | **Variant (NM_000138.5)** | **Primers** | **Sample** | **Laboratory** |
| --- | --- | --- | --- | --- |
| 1 | c.6872-11A>G | agaagaccgtaggatgtgca  tcctctgtcattgacacattc | Blood (PaxGene) | Wessex |
| 3 | c.6038-14T>G | gtagcgcgacggccagtACAGTGGGGTCTTTCCAGTG*  cagggcgcagcgatgacCCTCCTTCAAACTTCGCATA* | Blood (PaxGene) | Manchester |
| 4 | c.5671+773C>G | acctccacaggacagtgca  ccggcaagttccattccca | Blood (PaxGene) | Wessex |
| 6 | c.6617-8T>A | TGGAGGTTTTGAATGCACCT  TGCACATCCTACGGTCTTCT | Blood (PaxGene) | Wessex |
| 7 | c.1961-3T>G | ggattccagctggcatcag  tatcttatctgctacagtagag†  aagaagcatctgtcatcacac | Blood (PaxGene) | Wessex |
| 11 | c.3965-1081A>G | CGAATGCCGCATATCTCC  GATTTGCACTAATGCCTGACC  CACCTGTGTATCCTTCCTTGC  TGGGTTCAGTTCAAAATCAGG  TTCATTGATGTGAAGCCCAAT‡ | Blood (PaxGene) | Exeter |
| 12-15 | c.1589-1217G>T | ACCCCTGTGCTGGTGGTGAG CCCGCATTACACACGCAATG | Fibroblasts (+ve and -ve for cycloheximide) | From literature^2^ |
| 17 | c.1837+5G>C | ATTCATGCAGATCCCAGGGG  GGCCGGATCTGCAATAATGG | Blood (PaxGene) | Southampton |
| 20 | c.1469-244T>G | AGACTAACATTTCAGAAGTGGCT  CCAGTTGGTCCGCTATCTCT  GCATTCTGTCCGCGTGAG  CAGGGAATGCTTGGATATGGT | Fibroblasts | Oxford |
| 21 | c.6872-955C>G | tggaatgcaagaacctcattg  aaacccatcattacactcacag | Blood (PaxGene) | Wessex |
| 22 | c.6617-147G>A | tctgttggcaatccttgtgg  tgcacatcctacggtcttct | Blood (PaxGene) | Wessex |
| 23 | c.4942+19A>T | gatgagtgccaggagctac  gcaattatttcccccattca | Blood (PaxGene) | Wessex |
| 3, 5, 16 | c.6038-14T>G, c.164+96C>T, c.5422+452C>T | GCACCTTTGTGGTTCTCACT  GGGCCTAGTTGCAGTAGTTC | NA – primers for minigene assay | Manchester |

References

1.	Turnbull, C., Scott, R.H., Thomas, E., Jones, L., Murugaesu, N., Pretty, F.B., Halai, D., Baple, E., Craig, C., Hamblin, A., et al. (2018). The 100 000 Genomes Project: bringing whole genome sequencing to the NHS. BMJ *361*, k1687. 10.1136/bmj.k1687.

2.	Guo, D.C., Duan, X., Mimnagh, K., Cecchi, A.C., Marin, I.C., Yu, Y., Velasco, W.V., Lee, K., Zhu, X., Murdock, D.R., et al. (2023). An FBN1 deep intronic variant is associated with pseudoexon formation and a variable Marfan phenotype in a five generation family. Clin Genet *103*, 704-708. 10.1111/cge.14322.

3.	Guo, D.C., Gupta, P., Tran-Fadulu, V., Guidry, T.V., Leduc, M.S., Schaefer, F.V., and Milewicz, D.M. (2008). An FBN1 pseudoexon mutation in a patient with Marfan syndrome: confirmation of cryptic mutations leading to disease. J Hum Genet *53*, 1007-1011. 10.1007/s10038-008-0334-7.

4.	Gillis, E., Kempers, M., Salemink, S., Timmermans, J., Cheriex, E.C., Bekkers, S.C., Fransen, E., De Die-Smulders, C.E., Loeys, B.L., and Van Laer, L. (2014). An FBN1 deep intronic mutation in a familial case of Marfan syndrome: an explanation for genetically unsolved cases? Hum Mutat *35*, 571-574. 10.1002/humu.22540.

5.	Fusco, C., Morlino, S., Micale, L., Ferraris, A., Grammatico, P., and Castori, M. (2019). Characterization of Two Novel Intronic Variants Affecting Splicing in FBN1-Related Disorders. Genes (Basel) *10*. 10.3390/genes10060442.

6.	Torrado, M., Maneiro, E., Trujillo-Quintero, J.P., Evangelista, A., Mikhailov, A.T., and Monserrat, L. (2018). A Novel Heterozygous Intronic Mutation in the FBN1 Gene Contributes to FBN1 RNA Missplicing Events in the Marfan Syndrome. Biomed Res Int *2018*, 3536495. 10.1155/2018/3536495.

7.	Hiraide, T., Shimizu, K., Miyamoto, S., Aoto, K., Nakashima, M., Yamaguchi, T., Kosho, T., Ogata, T., and Saitsu, H. (2022). Genome sequencing and RNA sequencing of urinary cells reveal an intronic FBN1 variant causing aberrant splicing. J Hum Genet *67*, 387-392. 10.1038/s10038-022-01016-1.

8.	Kim, J.A., Jang, M.A., Jang, S.Y., Kim, D.K., Kim, Y.G., Kim, J.W., Park, T.K., and Jang, J.H. (2024). Overcoming challenges associated with identifying FBN1 deep intronic variants through whole-genome sequencing. J Clin Lab Anal *38*, e25009. 10.1002/jcla.25009.
